# Potential and Limitations of the MinION Nanopore array for miRNA-Enabled Early Cancer Detection

**DOI:** 10.1101/2025.02.18.25322511

**Authors:** Anastassia Kanavarioti, Aleena Rafiq

## Abstract

The 2024 Nobel Prize in Physiology or Medicine was awarded to the pioneers who reported that microRNAs (miRNAs) regulate and direct the switch between physiological and pathological pathways via their over- or under-expression. The discovery has not yielded any health benefit primarily due to raw data with no clear distinction between healthy and diseased blood samples. MiRNAs exist at femtomolar level in biological fluids, and typically quantified using amplification-based techniques. Experimental nanopores have illustrated potential for trace analysis including amplification-free miRNA quantification. We repurposed the MinION, the only commercially available nanopore-array device, and developed unique probes and protocols to quantify microRNAs in blood and urine. Our 2024 report revealed (i) miRNA copies are proportional to the total RNA isolated from the biospecimen, and (ii) several known miRNA cancer biomarkers were 1.8-fold overexpressed in blood samples from breast, prostate and pancreatic cancer patients compared to healthy. In contrast to literature reports sample variability was undetectable after normalization to the same RNA content. Here the earlier data were confirmed, and the conclusions extended to ovarian, lung, and colorectal cancer. The potential of the ready-to-use MinION/Yenos platform for multiple cancer early detection (MCED) using blood or urine is discussed.

## INTRODUCTION

By 2001 several lines of research had led to the discovery of miRNAs and the proposition that miRNAs control and regulate the post-transcriptional expression of proteins and the switch from physiological to disease pathways [1,2]. miRNAs are single-stranded (ss) RNAs around 22-nucleotide long, and stable in biological fluids which renders them suitable biomarkers [3,4]. Currently there are over 2,500 human miRNAs known [5,6], the subject of over 80,000 medical studies which have associated abnormal expression of selected miRNAs with cancer onset, progression, prognosis, and metastasis [7]. Due to the issues discussed below the potential of miRNAs as cancer biomarkers is at a standstill, and even responsible for delaying the development of anti-miRNA-based therapeutics [8,9]. Satisfactory resolution of these issues may revolutionize the miRNA field. Validation of selected miRNAs as early detection cancer biomarkers may lead to FDA approved miRNA-based liquid biopsies as screening/diagnostic or companion tests, minimize suffering, improve patient outcome, save lives, and reduce the cost of health care in our country and worldwide.

**Cancer screening** has been a major research front for decades [10-13]. The classical circulating blood biomarkers for cancer, such as Prostate-specific antigen (PSA), carcinoembryonic antigen (CEA), cancer antigen 125 (CA125), alpha-fetoprotein (AFP), etc., are neither sensitive nor specific. These tests are often inconclusive and are not recommended for population screening [12-13]. Several cancer indications, such as pancreatic, typically shows no early symptoms and late stage may be inoperable [14]. There is a world-wide push to include liquid biopsy tests as companion tests to help medical professionals cover the “grey” area of the current tests [15-20]. Most of the 80,000 “miRNA cancer” studies have concluded that a single miRNA or a miRNA panel may serve as cancer biomarker [7]. Typically, the data exhibit high variability and data from the cancer samples overlap with the data from the healthy samples. Investigators report the mean of the measurements and compare the mean from the cancer samples to the mean of the healthy samples [21-25]. Further statistical analysis yields a score of how well the specific miRNA, or miRNA panel prognoses cancer progression. A good score may be satisfactory in the case of a drug that treats a life-threatening disease, but a good score is not sufficient to diagnose cancer in an asymptomatic individual. Because of this “data spread”, no miRNA assay has received FDA approval as a screening test.

**The overlap of miRNA data between cancer and healthy samples** and the quantitative disagreement among studies have been attributed to differences in biospecimen collection methods, study protocols, choice of reference, analytical methods, population variation, age, sex, ethnicity, disease stage, etc. [21-25]. This uncertainty led to questioning the usefulness of miRNAs as liquid biopsy biomarkers. To the best of our knowledge, we were the first to report zero data overlap between healthy and cancer samples [26].

**The concentration of miRNAs in blood** is in the low femtomolar (fM) range, which is a billion-fold less than the micromolar (μM) range required by typical UV-Vis analytical tools. Current methods for assessing relative abundance of miRNAs in biological fluids or tissues depend mostly on amplification and include small RNA sequencing, reverse transcription-quantitative PCR (RT–PCR), droplet digital PCR (ddPCR), as well as microarray hybridization and Northern Blot [21-25]. These technologies require expensive infrastructure and skilled personnel, whereas our method only requires a laptop/internet, and could be implemented in a point-of-care (POC) facility [26]. Until now the PCR-based technologies were believed to be the most accurate; they typically report relative miRNA expression and not absolute copy numbers. Although identification works well with these tools, the quantification accuracy and choice of reference may be partially responsible for the “data spread”. A newer sequencing platform based on nanopore detection from Oxford Nanopore Technologies (ONT) introduced the MinION device 10 years ago and has been successfully applied to sequencing long DNA/RNA [27]. Recently the MinION’s sequencing approach was explored for multiplexed biomarker detection [28]. The authors reported miRNA detection limit (DL) at about 50 pM which is not suitable for liquid biopsy. For example, miR-16, one of the most abundant miRNAs in blood, measures about 170 fM [29], about 300-times less than the above DL. We also exploited the MinION platform but instead of sequencing we invented a direct detection/quantification (sensing) protocol, described earlier [26,29-31] and optimized further here. One of the least abundant miRNAs is miR-141 recently quantified at about 5 fM, a better than 10,000 sensitivity improvement over the MinION’s sequencing approach, illustrating the suitability of the MinION/Yenos platform for quantifying miRNAs in liquid biopsies [26].

## RESULTS

We hypothesized that miRNA quantification may be improved by developing an amplification-free nanopore array-based assay [30-33]. In 2020 we reported on the concept and implementation of a platform to measure single stranded nucleic acids, including miRNAs, in liquid biopsies (Fig. 1, [30-31]). In 2022 we posted the development and optimization of the MinION/Yenos platform [29]. In a 2024 report this technology showed discrimination between healthy and Stage I/II serum and urine samples from treatment-naïve patients with breast, prostate, and pancreatic cancer [26]. The latter included several conclusions, listed below, which were confirmed in this study and extended to Stage I/II serum samples from treatment-naïve patients with ovarian, colorectal and lung cancer. The technology yields miRNA copy number (miRNA #) per 1μL of total RNA isolate.

**Figure 1.**
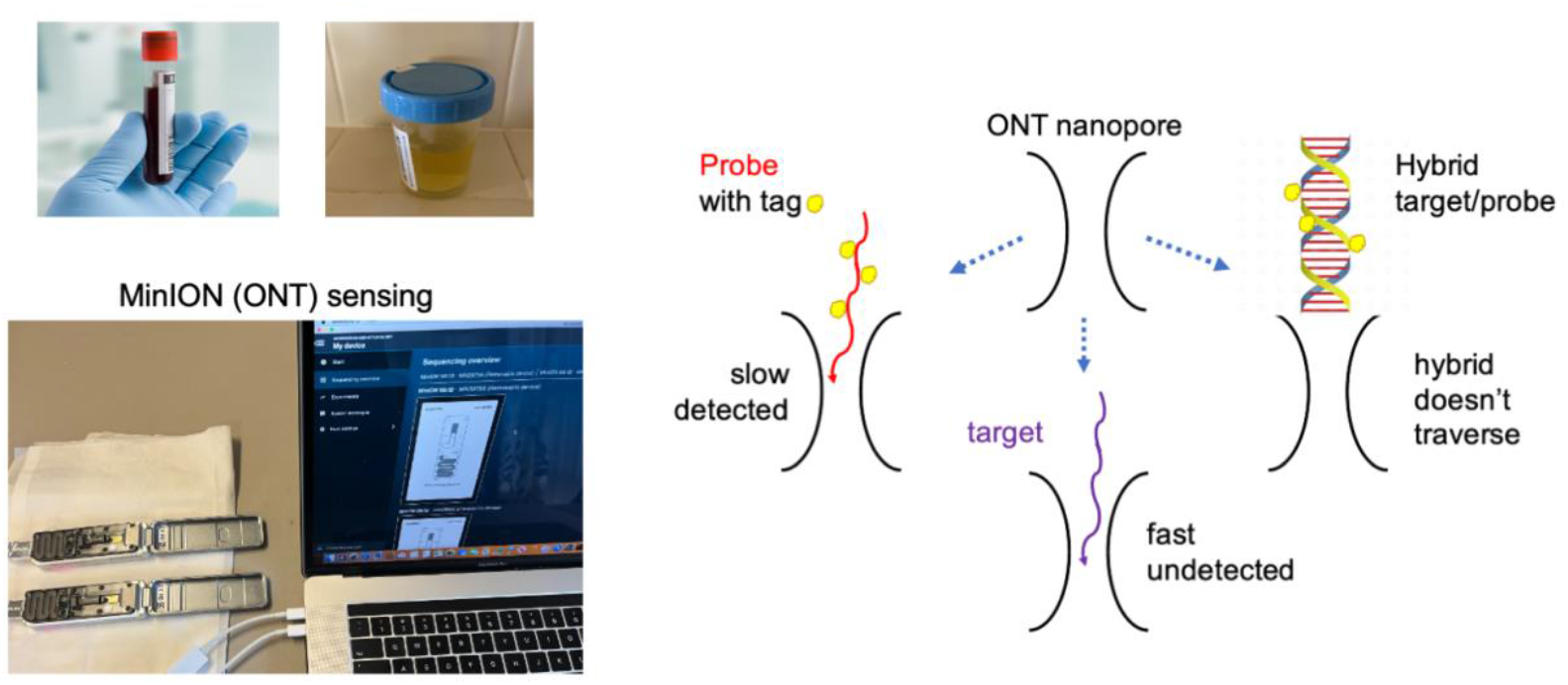
Graphical abstract of the processes involved in the miRNA measurement using the MinION platform. Left, top: Collection of the biospecimen, blood or urine, followed by total RNA isolation using a commercial kit and measurement of total RNA in the isolate using a nanodrop spectrophotometer. Left, bottom: Mixing an aliquot from the RNA isolate with an aliquot of the probe complementary to the target miRNA (Materials), adding ONT buffer and conducting a MinION ion conductance experiment (two experiments running simultaneously, shown here). The experiment measures the ion current (*I*) in picoamperes (pA) as a function of time (t) in milliseconds (ms). *I* is constant at *Io*, which is the open nanopore ion current (*Io*). When a single molecule traverses the nanopore, *Io* is reduced to a new value, *Ir*, because the molecule occupies the space that would have been occupied by the electrolyte that produces *Io*. Ion current reduction (dip in this platform) lasts for a time, τ and produces an event that is read by software (*OsBp_detect*, see Methods). The analysis determines whether the free probe is in excess and detected (left on the scheme above) or if the probe is not detected because it is hybridized with the target (right on the scheme above). Notably, RNAs, including the target miRNA, traverse much faster than the probes, and they are mostly missed (bottom on the scheme above) due to the relatively slow acquisition rate of this platform. A probe is an oligodeoxynucleotide complementary to the target miRNA, loaded with osmium tags (see Materials), optimized for hybridization with the target and for detection by the nanopores [29].

### Experimental Results Reported Earlier [26,29] and Confirmed in This Study

1. Most miRNA studies report relative expression. Mitchell et al. determined copy numbers for miR-15b and miR-16 from 3 healthy individuals and showed that miR-16 expression is approximately 10-fold higher than miR-15b [3]. These observations were confirmed quantitatively using our technology and the combined serum of healthy men, purchased from Millipore (Part No H6914) [29].
2. The Monarch New England Biolabs kit used to isolate total RNA, includes an additional protocol to isolate small RNA from the total RNA isolate. Independent measurements using the total RNA fraction or the small RNA fraction as the source of miR-15b gave comparable miR-15b #, suggesting that total RNA isolate is suitable for miRNA quantification and that the extra process of isolating small RNA from total RNA has no additional advantage [29].
3. According to Mitchell et al. miR-15b is not a cancer biomarker with comparable expression in samples from cancer patients or healthy individuals [3]. We confirmed this observation and were the first to report that miR-15b # depends on the total RNA content of the isolate (Fig. S1 in Supplementary Materials and [29]).
4. Over a period of 5 years, we purchased different lots of the combined serum product (H6914) and obtained comparable miRNA #. The different lots were analyzed by different analysts, using different protocols, different flow cell versions, and different probes, complementary to the target, but chemically distinct [29,26].
5. The 2024 report included a human study approved by the Advarra Investigational Review Board (IRB) on November 15, 2023. Protocol: Yenos Analytical LLC-02. “Quantification of selected microRNAs in the urine of healthy individuals” (Pro00074065) [26]. The study found that, after normalization to the same RNA content, miRNA # from urine samples of healthy subjects were all, within experimental error, comparable, as well as comparable to the corresponding miRNA in H6914 (see Fig. S2 in Supplementary Materials and [26]). In the IRB approved study total RNA in serum measured 6.9 < RNA < 16.4 ng/μL and in urine samples 8.7 < RNA <174.6 ng/μL. These observations implied that miRNA # are independent of age, sex, and ethnicity because the healthy subject cohort intentionally included subjects differing in age, sex, and ethnicity. The comparability between miRNA data from serum H6914 and healthy urine samples, suggested that, at least for the tested miRNAs, urine may replace blood draw.
6. Based on literature we anticipated that miRNA # in cancer samples would be many-fold elevated than in healthy samples [3]. This expectation didn’t materialize, and our preliminary observations pointed to a 2-to 3-fold overexpression [29]. Additional experiments and normalization to the same RNA content yielded a 1.8-fold overexpression (Fig. S2 in Supplementary Materials and [26]). A 1.8-fold overexpression requires measurement accuracy at +/-25% to achieve zero overlap between the data from the healthy and the cancer samples [26]. Our technology is based on a bracketing protocol designed to yield such accuracy and therefore it is well suited for these measurements. On the contrary, amplification methods may not be suitable to detect such a modest effect.
7. Figure S2 (Supplementary Materials) illustrates the zero data overlap and the 1.5 HL threshold between cancer samples and healthy samples [26]. The observed zero data overlap may not be limited to the small sample size. It is the product of the bracketing protocol, the exceptionally high accuracy imposed on the accepted measurement, and the suitable normalization approach (total RNA). Notably, all four tested biomarkers appear to be independent of the biospecimen, the collection method, age, sex, and ethnicity, and while three are cancer-depended, miR-15b is not.

### The current study

We manufactured new probes for each miRNA (miR-15b, miR-21, miR-375 and miR-141) based on published methods [26,29,30]. All the nanopore experiments reported here were conducted with the new probes. The reference which is the combined serum of healthy men (H6914 from Sigma Aldrich - Millipore, two new lots) was retested, and new serum samples were procured from Stage I/II, treatment-naïve patients with breast, prostate, pancreatic, ovarian, colorectal and lung cancers. Two healthy subjects provided urine samples. Total RNA was isolated from serum or urine using the appropriate commercial isolation kits (Methods and Materials). Cancer sera, H6914, and urine samples were used as a training set to optimize the experimental protocol of the nanopore experiments and the data analysis methodology. Selected experiments were conducted to show that cancer miRNA biomarkers are about 1.8-fold overexpressed and that miR-15b is not a cancer biomarker. This work focused on testing whether a certain miRNA measures above or below 1.5-fold of the reference (H6914 at 1.0 healthy level (HL) for all miRNAs). As miRNA # (1.0 HL) in all our studies we used the results obtained from our first study with miR-15b=17,710, miR-21=10,494, miR-375=9,240, and miR-141=6,096 per 1μL of total RNA or the equivalent of 2μL of serum [29]. Notably, the serum isolation kit yields 100μL of total RNA from 200μL of serum, and the urine isolation kit yields 50μL from 5mL of urine (Materials and Methods). Based on the earlier conclusions miR-15b must measure below the 1.5 HL threshold for all samples, whereas miR-21, miR-375, and miR-141 must measure above 1.5 HL in the cancer samples, but below 1.5 HL in the healthy samples (new lots of H6914 and the urine samples). Table 1 summarizes the results and the statistics for “true” result for each miRNA. Since the objective of the study was to optimize the protocol and the analysis, the data obtained are not all optimal. Presumably, the optimized protocol may produce statistics equal or better than obtained in this study.

**Table 1.** Testing for miRNA copies in samples (training set) at the 1.5 HL level of the reference H6914. Expectations: (i) miR-15b copies < 1.5 HL in all samples; miR-21, miR-375 and miR-141 > 1.5 HL in cancer samples, but < 1.5 HL in healthy samples. Measurement meeting expectation (ν), not meeting expectation (X) and not made (ND).

| **Cancer serum** | **ID** | **miR-15b** | **miR-21** | **miR-375** | **miR-141** |
| --- | --- | --- | --- | --- | --- |
| lung | A1 | v | v | v | v |
| ovarian | A2 | v | v | v | v |
| " | A3 | v | v | v | v |
| " | A4 | v | v | v | v |
| " | A5 | v | v | v | v |
| CRC<sup>1</sup> | No1 | v | v | v | v, v |
| lung | No2 | v, v | X | v, v | v, v |
| CRC<sup>1</sup> | No3 | v | ND | v | v, v, v |
| lung | No4 | v, v | v | v, v | v, v, v |
| lung | No6 | v | v, v | X | v, v, v |
| CRC<sup>1</sup> | No7 | v | v, v | v | v, v, v |
| prostate | D1 | v | v | v | X |
| pancreatic | D2 | v | v | X | v |
| breast | D3 | X | v | ND | v |
| " | D4 | v | v | v | v |
| prostate | D5 | X | v | v | ND |
| " | D6 | v | X | v | v, v |
| " | D7 | v | v | v | v |
| pancreatic | D8 | v | v | v | v |
| prostate | D9 | v | v | v | v |
| breast | D10 | v | v | X | v |
| pancreatic | D11 | v | v | X | v |
| **Healthy serum** | SQ H6914 | v, v, v | v, v, v | v, v | v, v |
| " | H6914 2025 | v | v, v | v | ND |
| **Healthy urine** | Healthy1 | ND | v, v | ND | v, v |
| " | Healthy2 | v, v | v, v | X | v |
| **Score** |  | *26/28* | *28/31* | *20/27* | *36/37* |
| **% True<sup>2</sup>** |  | **0.929** | **0.903** | **0.741** | **0.973** |

<sup>1</sup> CRC stands for colorectal. <sup>2</sup> % True for all 4 miRNAs with the 22 cancer samples stands at 0.912 and % True = 0.958 with the 4 healthy samples.

### The Potential and Limitations of the MinION/Yenos Platform for Cancer Screening

The data in Table 1 were collected with slightly modified protocols in search for an optimal one. The optimal protocol incorporates two 45 min experiments with buffer alone and two 45 min experiments with the sample; these four experiments represent a single “test”. Both the protocol to acquire the data and the methodology to obtain the result is detailed below. The sample to be tested contains a predetermined amount of probe that is 1.5-fold of the corresponding miRNA in the reference H6914, adjusted to the total RNA of the sample. The test has a binary result, namely the probe copies are either more or less than 1.5 HL. When the probe copies are more than the target miRNA, the result is detection of the free probe. When the probe copies are less that the target miRNA copies, then the result is no probe detection or silencing. Practically speaking the later experiment appears more like a buffer experiment.

This experimental approach targeting the 1.5 HL threshold works only because these three miRNA biomarkers were shown to be about 1.8-fold overexpressed in the cancer samples compared to healthy and because each one of the tested miRNAs is proportional to the total RNA of the sample. The accuracy of the optimized protocol is found to be comparable, at 97%, regardless of if the experiment is a detection or a silencing experiment. Earlier and current results support the proposition that each one of the 3 tested miRNA cancer biomarkers is a valid biomarker for each of the six cancer indications tested here. The different scores in Table 1 are attributed to experimenting with several less-than-optimal protocols.

Repeating the experiments with the optimal protocol would be time consuming and expensive and was done selectively for confirmation (multiple entries in Table 1). The Supplementary Materials contains the histogram, i.e., events plot vs. fractional residual ion current, of the test for each entry in Table 1.

**Assuming a 97% accuracy**, a single test per subject will yield 3 out of 100 false results. Two tests per subject will give 9 false results in a group of 10,000 subjects, and three tests will give 27 false results in a group of 1,000,000 subjects. These calculations presume that assignment will be made if all tests agree. Statistically, with 3 tests at a 97% accuracy, 8.7% will show agreement between two tests, while the third test will disagree. One way to reach a conclusion in such a case is to conduct a 4th test.

### Tentative Adaptation of the MinION to Sensing

Since the proposed protocol uses a “buffer only” experiment between samples, contamination is at a minimal and multiple samples can be analyzed in the same flow cell. However, the ONT proprietary flow cells are not resilient. This can be attributed to the proteinic nature of the nanopores and the fact that a nanopore is composed of 8 peptide strands that, to our knowledge, are not covalently attached to each other. If this is true, it is easy to see why the relatively fast translocation of millions of molecules reduces the pore’s lifetime. The friability of the nanopores reduces the number of active pores with use, and in turn, reduces the number of translocating molecules. The latter necessitates that a buffer experiment is conducted as control ahead of a new sample. A second issue is with the ONT buffer which may be optimal for sequencing but not for sensing (see Materials and Methods). Since the buffer is proprietary, it is only ONT that can optimize it for sensing. A third issue is that the flow cell is equipped with 2024 nanopores but there are only 512 detection channels, so that, perhaps randomly, different pores are selected every time a new experiment is initiated. In our experience, nanopores exhibit substantial differences among them and to partially equalize these differences the optimized protocol requires two buffer runs instead of one. Same number of detection channels and nanopores may be a better strategy for sensing. If ONT wishes to adapt their technology to sensing, they likely have more improvements at hand than the above recommendations. Currently one can conduct 3 such tests - each test consists of four 45 min nanopore experiments - on a single flow cell. Further optimization may be possible with the collaboration of Oxford Nanopore Technologies. A big step forward will be achieved when a comparable device is equipped with a flow cell made from solid state nanopores which should be more resilient than the proteinic nanopores. If the nanopores are wide enough to fit a single RNA strand but narrow enough to prohibit translocation of double stranded DNA/RNA, our probes should be usable without modification. Most importantly, the cost of a flow cell with a solid-state nanopore array may be much lower than the current flow cell cost.

## Discussion and Conclusions

Until recently sample variability has been the main culprit in the miRNA field prohibiting the exploitation of miRNAs as cancer/disease biomarkers in liquid biopsies. The MinION/Yenos platform described here and in earlier reports suggests that all healthy subjects, regardless of sex, age, and ethnicity, exhibit comparable miRNA copies for several miRNAs, including miR-15b which is not a cancer biomarker. This conclusion is borne out after normalizing miRNA copies to the same total RNA content (16.0 ng/μL used here) isolated from the sample. Total RNA from serum samples and urine samples is isolated using distinct isolation kits from two different manufacturers and still miRNA levels are comparable, indicating that miRNA levels are independent of isolation kit and method. It was proposed earlier and confirmed here that urine may replace blood at least for the miRNAs tested here and in the earlier studies [26,29]. Using the MinION/Yenos platform to directly measure femtomolar levels of single stranded nucleic acids, such as the miRNAs in liquid biopsies, reverses the belief that data variability is due to the sample and points to the analytical tools as suboptimal for nucleic acid trace quantification. Nevertheless, the qualitative conclusions from miRNA studies conducted using amplification techniques are most likely still valid and miRNAs are useful biomarkers [7].

Earlier studies using amplification techniques reported that miR-21, miR-375 and miR-141 are overexpressed in cancer sera compared to healthy sera [7,34,35]. We have confirmed this proposition and showed, for the first time to our knowledge, zero data overlap between cancer and healthy samples. Here we confirmed this proposition by testing for such 1.5-fold overexpression in six cancer indications. The data suggested that each of these 3 miRNAs may be exploited as a single cancer biomarker for breast, prostate, pancreatic, ovarian, colorectal, and lung cancers. Further experimentation may show whether (i) one of these miRNAs is more suitable than the others as a MCED biomarker, (ii) additional cancer indications exhibit the observed pattern and the 1.5 HL threshold with the three miRNAs tested above, (iii) additional miRNAs prove to be cancer biomarkers and (iv) other miRNAs prove to be biomarkers specific to a certain cancer indication and not agnostic as the three miRNAs tested here.

This work focused on optimizing the protocol to conduct and analyze the MinION experiments using the Yenos probes, while earlier work optimized the probe design and their hybridization efficiency with the miRNA target [26,29]. Earlier work also explored multiplexing of probes, which is possible and perhaps more useful in cases where the overexpression of a miRNA in a cancer is higher than 2-fold. It is proposed that the optimized methodology will yield 97% accuracy per test regardless of if the test yields miRNA overexpression or not. If future work confirms the 97% accuracy, then testing a subject by conducting 3 such tests can be done with a single flow cell and should yield the “false” assignment in about 30 subjects in a million. Considering that our studies included treatment-naïve subjects, it is proposed that the MinION/Yenos test, as optimized here, may be used as a companion test to facilitate doctor/patient decisions when the current tests for breast, prostate, pancreatic, ovarian, colorectal and lung cancer fall into the “grey area”. Two MinION/Yenos tests may be sufficient for such a companion test. Implementation in the general asymptomatic population will need to wait until there is evidence that miRNA cancer biomarkers are overexpressed years ahead of any symptoms. The latter study may be conducted using the Prostate, Lung, Colorectal, and Ovarian (PLCO) samples collected by the National Cancer Institute (NCI) from asymptomatic volunteers who developed cancer years after blood draw.

## Materials and Methods

### Human Samples

Human serum from the USA, isolated via sterile filtration from male AB-clotted whole blood (H6914) was purchased from Sigma-Aldrich (Millipore now) and has been used as the reference for all our work. Different lots have been purchased and tested over the last 5 years and comparable results have been obtained. In all the studies miRNA # at 1.0 HL are the miRNA # determined in the first such study [29]. New lots used in this study are SLCN9218 (SQ H6914) and 0000305265 (H6914 2025 or H6914 12.1ng/μL). Serum samples purchased from Discovery Life Sciences (DLS, Huntsville, AL, USA) and Tissue for Research (Accio Biobank online, Suffolk, UK) were collected from informed consented individuals under the IRB/EC protocol. The selection of these samples from a large depository included both male and female donors, if applicable, and one each from African American, Hispanic, or White ethnicity. Samples were collected from newly diagnosed, naïve patients ahead of treatment. The demographics of the patients with cancer who provided their specimens is listed in Table S1 (Supplementary Materials). The project to include urine samples collected by themselves at home and sent by FedEx overnight to our facilities from consenting healthy subjects was reviewed by the Advarra Investigational Review Board (IRB). The protocol and consent form were reviewed, modified, and approved by the Advarra IRB on Nov. 15, 2023. Protocol: Yenos Analytical LLC-02. Quantification of selected microRNAs in the urine of healthy individuals (Pro00074065), with continuing review approval on 25 Oct. 2024 (CR00599062). Donors of urine samples reviewed and signed an informed consent form. For the isolation of total RNA from serum, we used the Monarch T2010S Kit from New England Biolabs. For the isolation of total RNA from urine, Norgen Biotek Corp. No. 29600 kit was used with 5 mL urine sample. Kits were used according to the manufacturer’s instructions.

### Oligos, Probes, and Other Reagents

The only ONT kit used for the experiments reported here was the Flow Cell Priming Kit XL (EXP-FLP002-XL), ONT flush buffer or ONT buffer. The ONT buffer is proprietary, provides the necessary electrolytes and must represent more than 80% of the 80-85 μL sample volume. miRNAs, custom-made DNA oligos and 2’-OMe-oligos, synthesized at the 0.1-0.25 μmole scale and purified by HPLC or PAGE by the manufacturer, were purchased from Integrated DNA Technologies (IDT). Oligos (sequences in Table 2) were diluted with Ambion nuclease-free water, untreated with DEPC, typically to 100 or 200 μM stock solutions, and stored at −20°C. The oligo purity was confirmed to be >85% by in-house HPLC analysis [36]. Osmylated oligos were manufactured in-house, based on published methods at a concentration of approximately 20 μM [see below] and purified using kit QIAquick Nucleotide Removal Kit cat. no. 28306 from Qiagen. Following osmium tagging (osmylation, see below), in-house HPLC analysis was used to determine the probe content, extent of osmylation, and efficiency of probe/target hybridization [29]. Osmylated oligos are stable and can be kept at -20°C for 2 years. LoBind Eppendorf test tubes (1.5 mL) were used for serial 5/1000 or 10/1000 dilutions to yield probes at concentrations at about 40 fM. Mixtures of aliquots from the probe and from the isolated RNA were prepared in 0.5 mL RNase-, DNase-free, sterile test tubes and used after addition of 75 μL of ONT buffer.

**Table 2.**
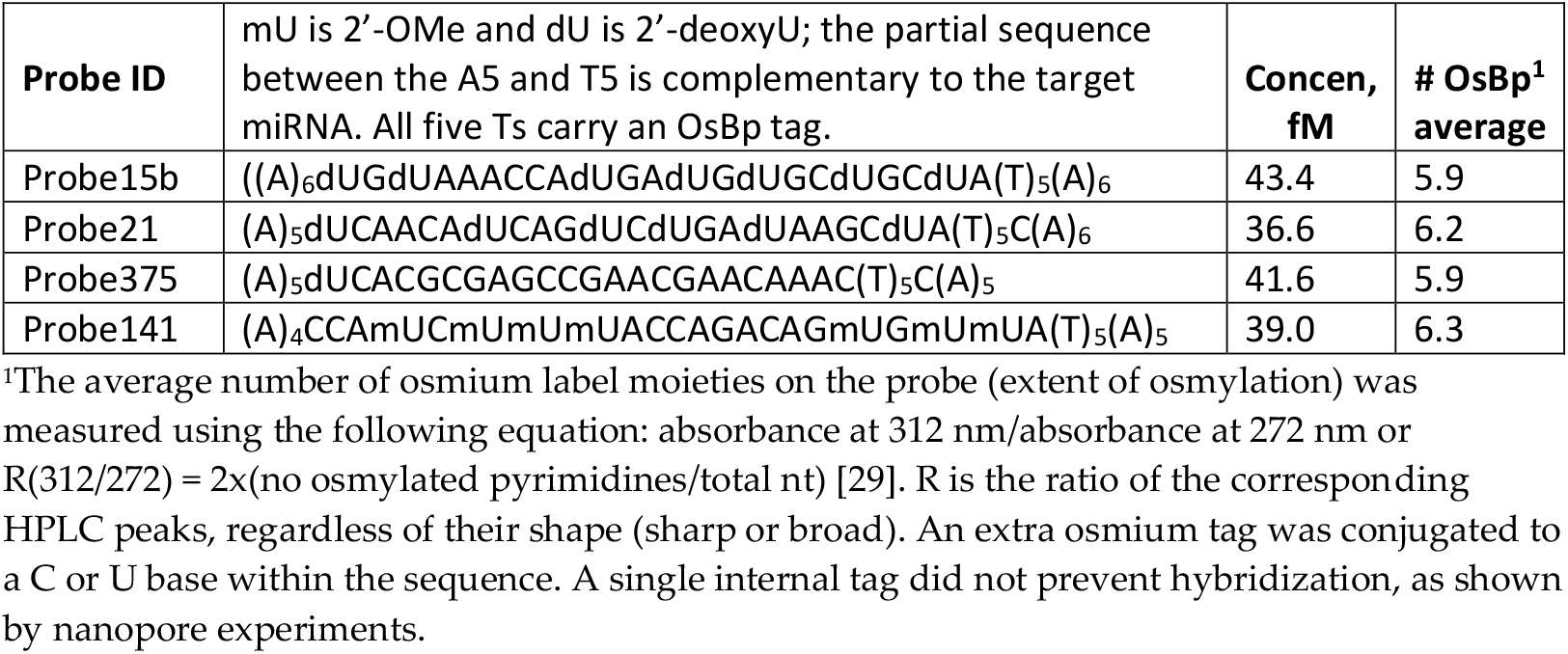
Sequence and characterization of the probes used in this work.

The probes were developed and optimized earlier [29-31]. They are oligodeoxynucleotides with a sequence complementary to the target miRNA but extended at one end with five adjacent thymidine (T) residues and flanked by up to five adenosines (A) at either end (Table 2). The A-tails facilitated the entry of the probe into the nanopores. The adjacent Ts were each tagged with an osmium label, i.e., osmylated [37-43] with a 1:1 mixture of OsO_4_ and 2,2′-bipyridine (abbreviated OsBp), to slow down their translocation via the nanopores and discriminate them from native nucleic acids. Within the probe sequence complementary to the target, Ts were replaced by uridine (U), 2’-OMe-U or dU to minimize OsBp labeling because the osmylation kinetics of U and cytosine (C) are substantially slower than that of T [38,40,41]. HPLC analysis yields the probe’s concentration (content) using intact oligo as a standard because the absorbance of the probe at 260 nm is practically the same as that of the precursor intact oligo [38]. HPLC analysis provided evidence of the quantitative depletion of the OsBp reagent which elutes ahead of the osmylated oligo.

### Sample Preparation

This technology determines miRNA copy number by bracketing. At least two measurements are necessary and typically more than two are needed for quantification. The spread between the measurements determines the accuracy; for technical reasons accuracy at +/-20% is a lower limit. Probe copies matter and should be kept around 100,000, somewhat more when the flow cell is new and somewhat less when the flow cell is old. The expected free probe copies should be detectable over the noise observed at the (*Ir/Io*)_max_ =0.15. A spreadsheet may be used to calculate simultaneously the amount of total RNA sample and probe to target miR-15b at 1.56 HL (Table 3). If the Yenos test (4 experiments) is judged to be detection, then probe > target and if silencing then probe < target, or the equivalent target < 1.56 HL or target > 1.56 HL, respectively.

**Table 3.** Sample preparation to detect miR-15b at the 1.56 HL in a sample of total RNA at 12.1 ng/μL using 4μL of 43.4 fM probe15b.

|  |  |  |  |
| --- | --- | --- | --- |
| Sample, μL | 5 |  |  |
| Probe15b copies | Probe15b copies per 1μL sample | Probe15b copies per 1μL sample normalized to reference at 16 ng/μL | Probe15b copies per 1μL sample normalized to the reference at 16 ng/μL and adjusted to target 1.56 HL miR-15b at 17,710 copies per 1μL RNA from H6914 reference |
| = 4*43.4*600<sup>1</sup> | = 4*43.4*600/5 | = 4*43.4*600/5*16/12.1 | = 4*43.4*600/5*16/12.1/17710 = 1.56 |

<sup>1</sup> 4x43.4x600 comes from 4x10<sup>-6</sup>x43.4x10<sup>-15</sup>x6.0x10<sup>23</sup>

### Single-Molecule Ion-Channel Conductance Experiments on the MinION (MinION Mk1B Platform)

Registration is required at the ONT website to download the software MinKNOW to a computer/laptop with specifications provided by ONT. All the functions necessary to test the hardware and flow cells and run the experiments were performed using the MinKNOW software. The sample was loaded onto a flow cell fitted in a MinION device. The experiment was run under “start sequencing” mode. A direct RNA sequencing kit (SQK-RNA002) was selected to initiate experiments. The flow cell type FLO-MIN106 was selected, and the run length (45 min) and bias voltage (-180 mV) were selected; basecalling was disabled, and the output bulk file Raw (1-512) was checked and generated. The output location was Library/MinKNOW/data/, and the output format was *fast-5*. All the experiments reported here were run for 45 min at -180 mV. This type of sensing experiment produces less than a million of selected (by our algorithm) events in 45 min. 45 min was used to permit the use of Microsoft Excel which limits the reported rows to about a million entries. The *fast-5* file was analyzed using the *OsBp_detect* algorithm [31]. The number of events per channel from the *OsBp_detect* analysis has been compared with the actual *i-t* trace of the specific channel using MATLAB visualization, and this algorithm, 2^nd^ revision, has been repeatedly validated. Currently, *OsBp_detect* can only be used with a 2017 or an earlier version of MacBook Pro loaded with macOS 10.14 Mojave. While alternative parameters were explored earlier, all experiments reported here were analyzed using the following threshold parameters: (i) event duration (in tps): 4-1200 (1.3-400 ms), (ii) lowest *I*_*r*_*/I*_*o*_ <0.55, and (iii) all *I*_*r*_*/I*_*o*_ <0.6, channels 1-512.

### Data Analysis

A laptop/computer requires a few min for the *OsBp_detect* analysis of the *fast-5* file, produces a file in *tsv* format, to be opened via Microsoft Excel and saved as such. In the Excel spreadsheet, the algorithm-selected events (*I*_*r*_*/I*_*o*_ data) are grouped in the form of a histogram with 0.05 bins, from 0.05 to 0.55, and plotted (Supplementary Materials, pages 4-22). Histograms exhibit two maxima *(I*_*r*_*/I*_*o*_)_*max*_: an early one at *I*_*r*_*/I*_*o*_ =0.15 and a late one at *I*_*r*_*/I*_*o*_ =0.30. The Yenos probes translocate preferentially at the early (*I*_*r*_*/I*_*o*_)_*max*_ =0.15. These maxima may vary by ±0.05 units depending on the flow cell age. The events under late *(I*_*r*_*/I*_*o*_)_*max*_ and early *(I*_*r*_*/I*_*o*_)_*max*_ were noted, and their *ratio R = ((I*_*r*_*/I*_*o*_)_*max*_ *late (0*.*3)/early (0*.*15))* was calculated. R value represents the criterion by which an experiment is judged as detection or silencing compared to the buffer control. The number of events decreases with flow cell use, but R remains comparable in the absence of free probe. In comparing sample to buffer run, a decreasing *ratio* R indicates detection (free probe present), and a comparable or increasing *ratio* R indicates silencing (no free probe present). Comparison of R values is deemed significant at more than 18% change. The 18% change is not set in stone, and confirmation can be done by running the sample again (see “nonew2” or 3^rd^ run in the Supplementary Materials, e.g., pages 12, 21 and 21). This assignment is consistent with an increased number of events owing to the presence of the probe, which traverses with *(I*_*r*_*/I*_*o*_)_*max*_ ∼ 0.15, whereas intact RNA and background noise traverse mostly with *(I*_*r*_*/I*_*o*_)_*max*_ ∼ 0.30. Buffer was run twice, and sample was run twice, and R values are calculated for each. Buffer and sample data may be added together and reported as 1.5 h (see Supplementary Materials, pages 4-21).

Because the flow cells lost active pores during every experiment, the *total* number of events decreases constantly with use. More than 200 active pores yield reliable data. Data analysis was optimized by deleting all channels which report more than 20,000 events; typically, these channels represent less than 5%. Reasons to reject a test are when (i) one sample run exhibited an R value increasing and the other sample run exhibited an R value decreasing compared to buffer R value, (ii) the *(I*_*r*_*/I*_*o*_)_*max*_ shifted by more than ±0.05 units or (iii) the sample produced events in large excess over the events produced by the buffer (see page 22 in Supplementary Materials).

## Data Availability

All data produced in the present study are available upon reasonable request to the authors

## Supplementary Materials

Page 1, list of figures and tables. Page 2, earlier work Figures S1 and S2. Page 3, Table S1: Demographics of patients included in Table 1 (text). Pages 4-21, Figures with nanopore tests to target the 1.5 HL H6914 threshold with miR-15b, miR-21, miR-375 and miR-141. Page 22, Rejected tests due to unacceptable event profiles.

## Author Contribution

Conceptualization, methodology, software, formal analysis, investigation, resources, and writing, A.K.; investigation, A.R. Both authors have read and agreed to the published version of the manuscript.

## Institutional Review Board Statement

The study was conducted in accordance with the Declaration of Helsinki and approved by the Advarra Institutional Review Board on 15 November 2023 (Protocol Pro00074065 entitled Quantification of selected microRNAs in the urine of healthy individuals) with continuing review approval on 25 October 2024 (CR00599062).

## Informed Consent Statement

Informed consent was obtained from all subjects involved in the study.

## Data Availability Statement

The data generated during this study are included in this published article. Raw data (fast5 format at ∼3.3 GB each) may be obtained from the corresponding author.

