## Supplemental Materials for "Potential and Limitations of the MinION Nanopore array for miRNA-Enabled Early Cancer Detection" (from another version of this preprint: version 1, 2025-02-21)

Page 2: Earlier work: Figures S1 and S2
Page 3: Table S1: Demographics of patients included in Table 1 (text)
Pages 4-22: Figures with MinION/Yenos tests to target the 1.5 HL H6914 threshold with miR-15b, miR-21, miR-375 and miR-141. Event plots of selected events (y-axis) grouped in 0.05 bins with $0 < Ir/Io < 0.55$ (x-axis)
Page 4: Nanopore experiments with samples ID (A1 and A2)
Page 5: Nanopore experiments with samples ID (A3 and A4)
Page 6: Nanopore experiments with sample ID A5
Page 7: Nanopore experiments with sample ID No1,
and evidence that 1.50 HL < No1 CRC miR-141 < 2.00 HL
Page 8: Nanopore experiments with sample ID No2, and
evidence that 0.72 HL < No2 lung miR-15b < 1.46 HL and
1.52 HL < No2 lung miR-375 < 2.08 HL
Page 9: Nanopore experiments with sample ID No3
Page 10: Nanopore experiments with sample ID No4 and
evidence that 1.58 HL < No4 lung miR-375 < 2.05 HL and
1.56 HL < No4 lung miR-141 < 2.05 HL
Page 11: Nanopore experiments with sample ID No6
Page 12: Nanopore experiments with sample ID No7
Page 13: Nanopore experiments with samples ID (D1 and D2)
Page 14: Nanopore experiments with samples ID (D3 and D4)
Page 15: Nanopore experiments with samples ID (D5 and D6)
Page 16: Nanopore experiments with samples ID (D7 and D8)
Page 17: Nanopore experiments with samples ID (D9 and D10)
Page 18: Nanopore experiments with sample ID D11
Page 19, 20: Nanopore experiments with samples ID SQ H6914 and H6914 2025 and
Evidence that 0.69 HL < SQ H6914 miR-15b < 1.52 HL and
0.69 HL < SQ H6914 miR-21 < 1.53 HL and
0.74 HL < H6914 (12.1ng/µL) miR-21 < 1.52 HL
Page 21: Nanopore experiments with samples ID Healthy1 urine and Healthy2 urine with
0.70 HL < Healthy1 urine miR-21 < 1.54 HL and
0.70 HL < Healthy1 urine miR-141 < 1.53 HL and
0.71 HL < Healthy2 urine miR-15b < 1.50 HL and
0.73 HL < Healthy2 urine miR-21 < 1.50 HL.
Page 22: Representative experiments which don’t pass quality control and are therefore rejected.

Earlier work included in Reference 26. Both figures also included in a poster presented at the AACR special conference in cancer research: Liquid biopsy: From discovery to clinical implementation held on Nov 13-16, 2024 in San Diego, CA, USA

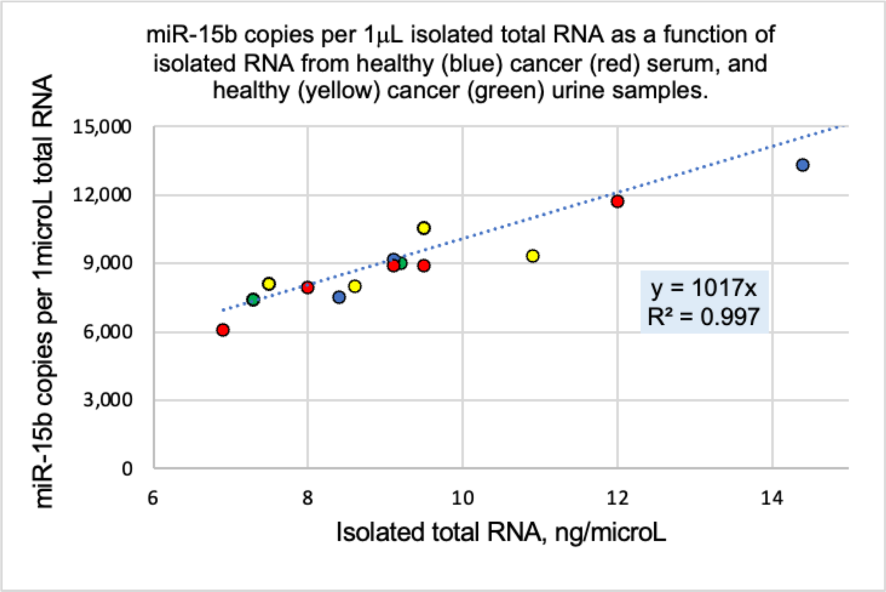


**Figure S1:** miR-15b copies are proportional to the total RNA content of the sample isolated from the biospecimen, and independent of source (blood or urine), RNA collection kit, age, sex, ethnicity, and the presence/absence of cancer.

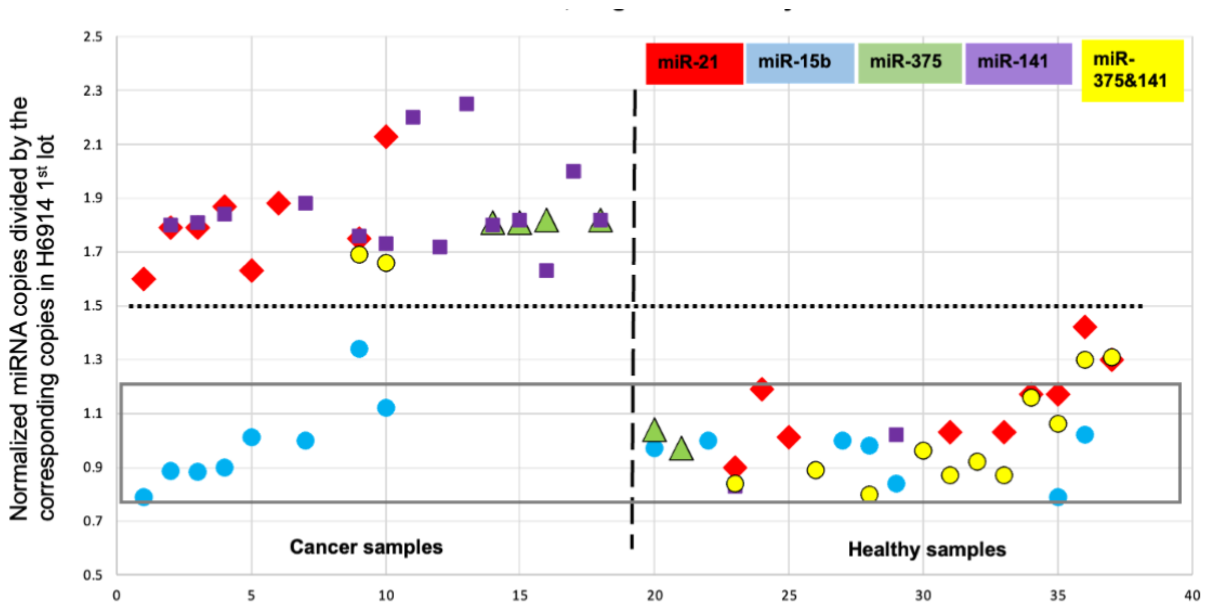


**Figure S2**: miRNA copies were normalized to the same RNA content (16 ng/µL) and then divided by the corresponding miRNA copy number in the reference H6914 (combined serum, healthy men assigned as 1.0 HL) to obtain x-fold HL (HL stands for healthy level). In cancer serum and urine samples miR-21, miR-375, miR-141 measured 1.83 HL (RSD=0.09), and in healthy urine samples 1.04 HL (RSD=0.17) with zero data overlap, p-value 1.6 x 10-[22]. Threshold between cancer and healthy samples is at 1.5 HL (dotted line).

**Table S1:** Demographics of cancer patients whose serum samples are listed in Table 1

| ID | Age group | Gender | Cancer | Stage, Note1 |
|---|---|---|---|---|
| A1 | 71-75 | M | lung | pT1c, pN0 |
| A2 | 46-50 | F | ovarian | pT1b, pN |
| A3 | 66-70 | F | ovarian | pT1a, pN0 |
| A4 | 56-60 | F | ovarian | pT1a, pN0 |
| A5 | 71-75 | F | ovarian | pT1c2, pN0 |
| No1 | 61-65 | F | Colorectal Cancer | Note1 |
| No2 | 61-65 | F | Lung Cancer, Adenocarcinoma | " |
| No3 | 76-80 | M | Colorectal Cancer, Adenocarcinoma | " |
| No4 | 66-70 | M | Lung Cancer, Squamous Cell Carcinoma | " |
| No6[1] | 66-70 | M | Lung Cancer, Squamous Cell Carcinoma | " |
| No7 | 51-55 | F | Colorectal Cancer, Adenocarcinoma | " |
| D1 | 41-45 | F | Breast Cancer, Ductal Carcinoma | I-A |
| D2 | 66-70 | F | Pancreatic Cancer, Adenocarcinoma | I-B |
| D3 | 41-45 | F | Breast Cancer, Ductal Carcinoma | II-B |
| D4 | 46-50 | F | Breast Cancer, Ductal Carcinoma | I-A |
| D5 | 71-75 | M | Prostate Cancer, Adenocarcinoma | I |
| D6 | 76-80 | M | Prostate Cancer, Adenocarcinoma | I |
| D7 | 56-60 | M | Prostate Cancer, Adenocarcinoma | I |
| D8 | 66-70 | M | Pancreatic Cancer | I-B |
| D9 | 56-60 | M | Prostate Cancer, Adenocarcinoma | II-C |
| D10 | 66-70 | F | Breast Cancer, Ductal Carcinoma | II-A |
| D11 | 66-70 | M | Pancreatic Cancer, Progressive/Refractory/Relapsed | II-A |

[1] No5 was a serum sample that exhibited total RNA at about 9 ng/μL and was not tested because the RNA content is at the lower limit of the assay.

Note 1: All patients were consented. Except for patient D11, all patients were newly diagnosed and treatment naïve. Patient ID No5 was not tested because total RNA measured 9ng/μL which is a relatively low level of total RNA and may not lead to the desired accuracy of +/-25%.

Group with samples ID A1-A5 obtained from Corewell Health West Biobank, 35 Michigan str. NE, MC056, 6th floor, Rm 6414, Grand Rapids, MI 49503. Arranged by Accio Biobank Online (Tissue For Research) www.biobankonline.com

Groups with samples ID No1-No7 and ID D1-D11 obtained from Discovery Life Sciences, 900 Hudson Way, Huntsville, AL 35806 http://www.dls.com.

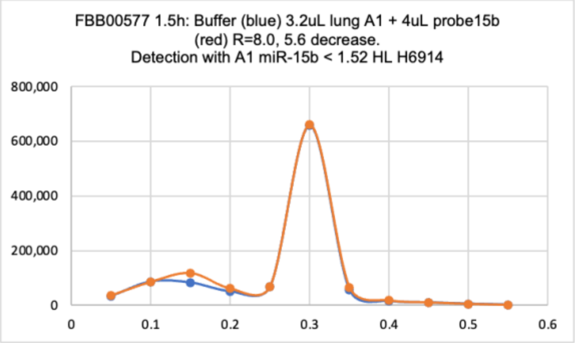
FBB00577 1.5h: Buffer (blue) 3.2uL lung A1 + 4uL probe15b (red) R=8.0, 5.6 decrease.
Detection with A1 miR-15b < 1.52 HL H6914
800,000
600,000
400,000
200,000
0
0 0.1 0.2 0.3 0.4 0.5 0.6

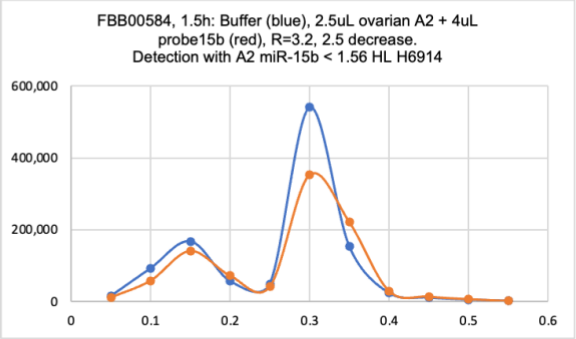
FBB00584, 1.5h: Buffer (blue), 2.5uL ovarian A2 + 4uL probe15b (red), R=3.2, 2.5 decrease.
Detection with A2 miR-15b < 1.56 HL H6914
600,000
400,000
200,000
0
0 0.1 0.2 0.3 0.4 0.5 0.6

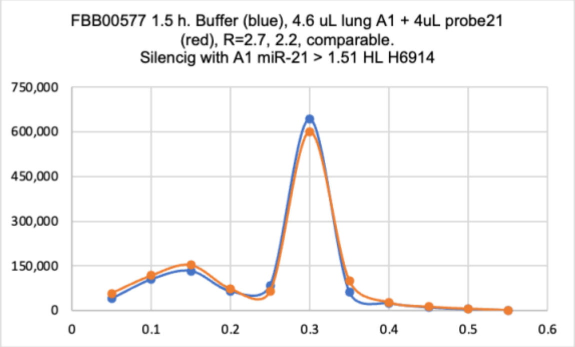
FBB00577 1.5 h. Buffer (blue), 4.6 uL lung A1 + 4uL probe21 (red), R=2.7, 2.2, comparable.
Silencig with A1 miR-21 > 1.51 HL H6914
750,000
600,000
450,000
300,000
150,000
0
0 0.1 0.2 0.3 0.4 0.5 0.6

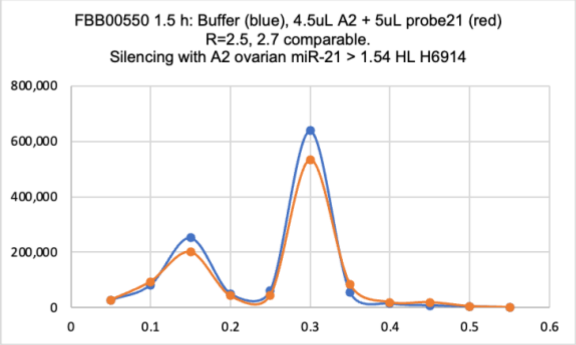
FBB00550 1.5 h: Buffer (blue), 4.5uL A2 + 5uL probe21 (red) R=2.5, 2.7 comparable.
Silencing with A2 ovarian miR-21 > 1.54 HL H6914
800,000
600,000
400,000
200,000
0
0 0.1 0.2 0.3 0.4 0.5 0.6

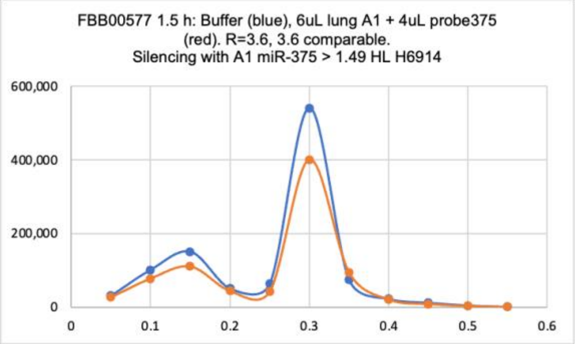
FBB00577 1.5 h: Buffer (blue), 6uL lung A1 + 4uL probe375 (red). R=3.6, 3.6 comparable.
Silencing with A1 miR-375 > 1.49 HL H6914
600,000
400,000
200,000
0
0 0.1 0.2 0.3 0.4 0.5 0.6

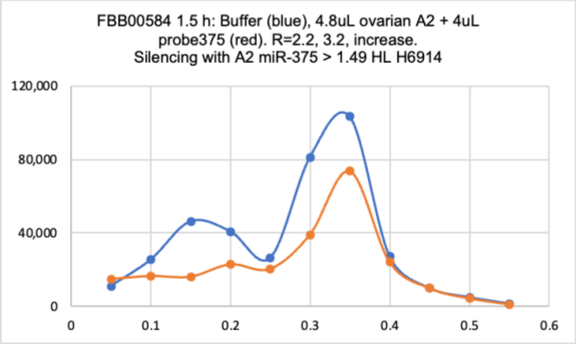
FBB00584 1.5 h: Buffer (blue), 4.8uL ovarian A2 + 4uL probe375 (red). R=2.2, 3.2, increase.
Silencing with A2 miR-375 > 1.49 HL H6914
120,000
80,000
40,000
0
0 0.1 0.2 0.3 0.4 0.5 0.6

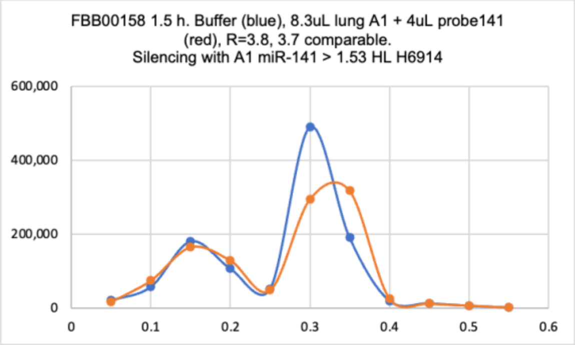
FBB00158 1.5 h. Buffer (blue), 8.3uL lung A1 + 4uL probe141 (red), R=3.8, 3.7 comparable.
Silencing with A1 miR-141 > 1.53 HL H6914
600,000
400,000
200,000
0
0 0.1 0.2 0.3 0.4 0.5 0.6

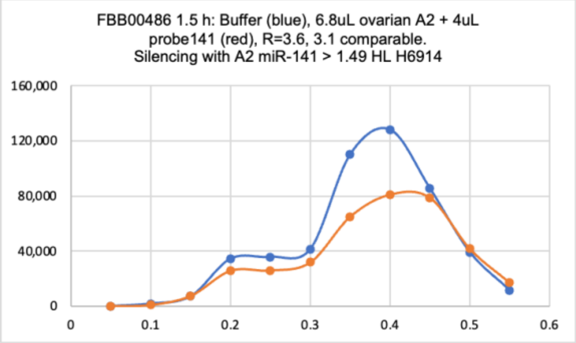
FBB00486 1.5 h: Buffer (blue), 6.8uL ovarian A2 + 4uL probe141 (red), R=3.6, 3.1 comparable.
Silencing with A2 miR-141 > 1.49 HL H6914
160,000
120,000
80,000
40,000
0
0 0.1 0.2 0.3 0.4 0.5 0.6

FBB00661, 1.5 h: Buffer (blue), 3uL ovarian A3 + 4uL probe15b (red), R=3.2, 2.2 decrease.
Detection with A3 miR-15b < 1.48 HL H6914

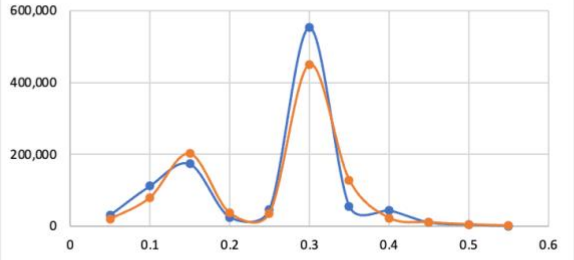


FBB10401 1.5 h: Buffer (blue), 5.2uL A3 (ovarian) + 5uL probe21 (red), R=2.7, 2.7 comparable.
Silencing with A3 miR-21 > 1.52 HL H6914

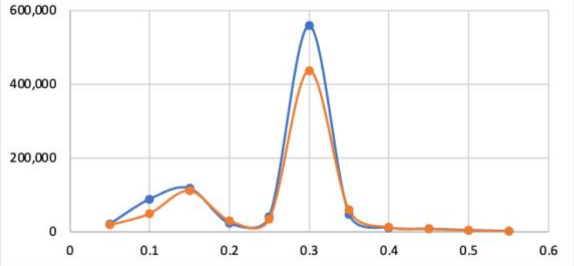


FBB00661 1.5 h. Buffer (blue), 5.4uL ovarian A3 + 4uL probe375 (red), R=2.1, 1.9 comparable.
Silencing with A3 miR-375 > 1.51 HL H6914

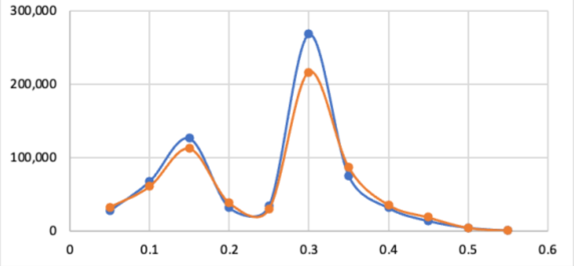


FBB00503 1.5 h: Buffer (blue), 7.6uL ovarian A3 + 4uL probe141 (red), R=3.7, 3.2 comparable.
Silencing with A3 miR-141 > 1.52 HL H6914

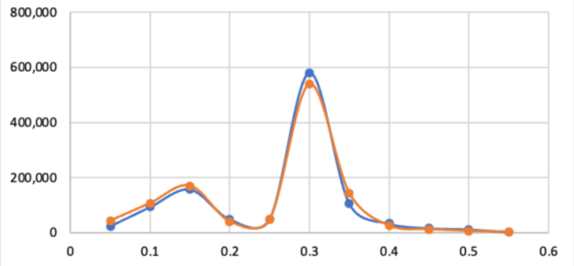


FBB10383 1.5h: Buffer (blue), 3.9uL ovarian A4 + 4uL probe15b (red). R=3.1, 2.5, decrease.
Detection with A4 miR-15b <1.51 HL H6914

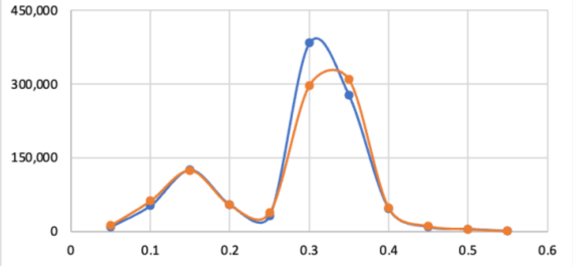


FBB10383 1.5h Buffer (blue), 5.5uL ovarian A4 + 4uL probe21 (red), R=2.5, 2.9 comparable.
Silencing with A4 miR-21 > 1.52 HL H6914

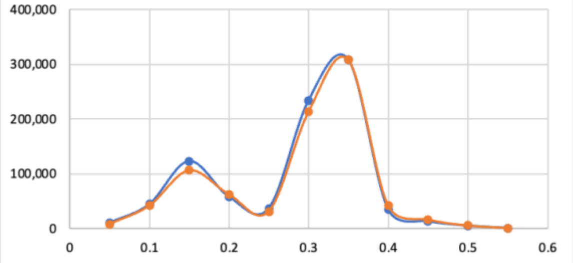


FBB10383 1.5 h. Buffer (blue), 7.2uL ovarian A4 + 4uL probe375 (red) R=2.0, 2.1 comparable.
Silencing with A4 miR-375 > 1.50 HL H6914

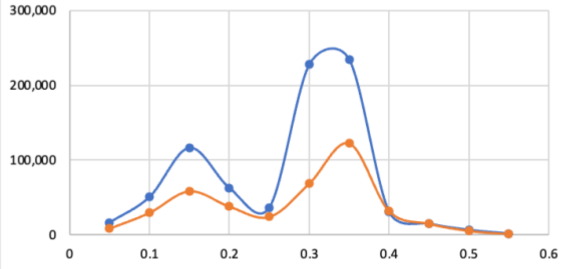


FBB00596 45min. Buffer (blue), 10uL ovarian A4 + 4uL probe141 (red) nonew (green), R=2.5, 2.4, 2.0 comparable.
Silencing with A4 miR-141 > 1.54 HL H6914

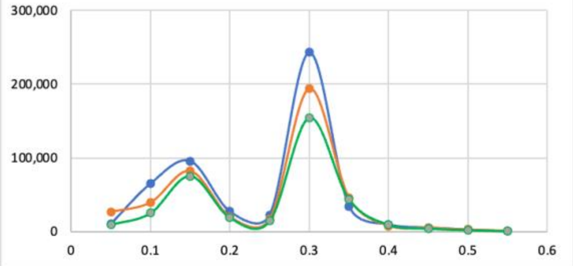

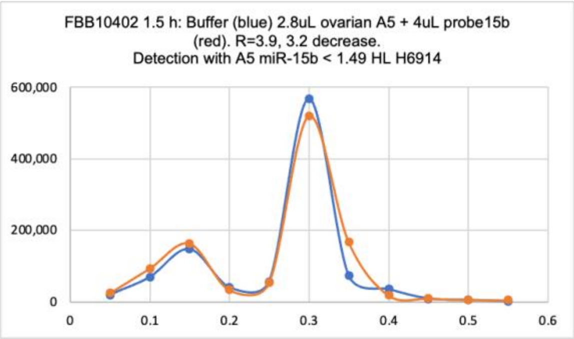

FBB10402 1.5 h: Buffer (blue) 2.8uL ovarian A5 + 4uL probe15b (red). R=3.9, 3.2 decrease.
Detection with A5 miR-15b < 1.49 HL H6914

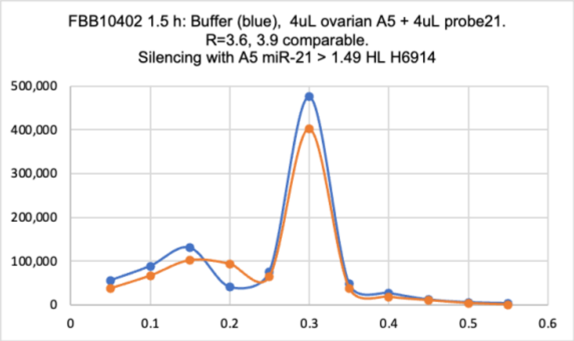

FBB10402 1.5 h: Buffer (blue), 4uL ovarian A5 + 4uL probe21. R=3.6, 3.9 comparable.
Silencing with A5 miR-21 > 1.49 HL H6914

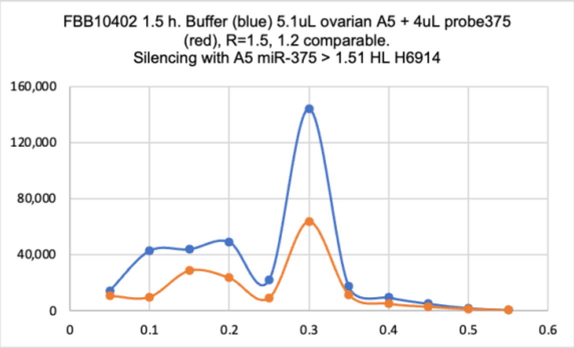

FBB10402 1.5 h. Buffer (blue) 5.1uL ovarian A5 + 4uL probe375 (red), R=1.5, 1.2 comparable.
Silencing with A5 miR-375 > 1.51 HL H6914

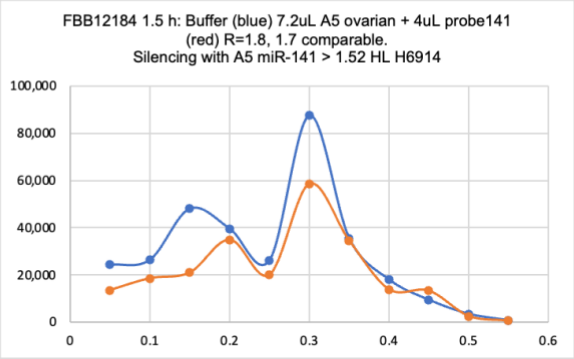

FBB12184 1.5 h: Buffer (blue) 7.2uL A5 ovarian + 4uL probe141 (red) R=1.8, 1.7 comparable.
Silencing with A5 miR-141 > 1.52 HL H6914

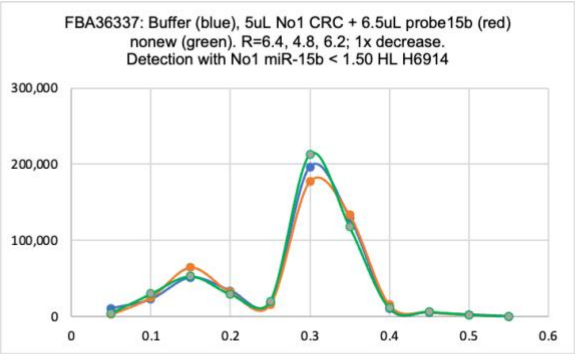
FBA36337: Buffer (blue), 5uL No1 CRC + 6.5uL probe15b (red)
nonew (green). R=6.4, 4.8, 6.2; 1x decrease.
Detection with No1 miR-15b < 1.50 HL H6914
300,000
200,000
100,000
0
0 0.1 0.2 0.3 0.4 0.5 0.6

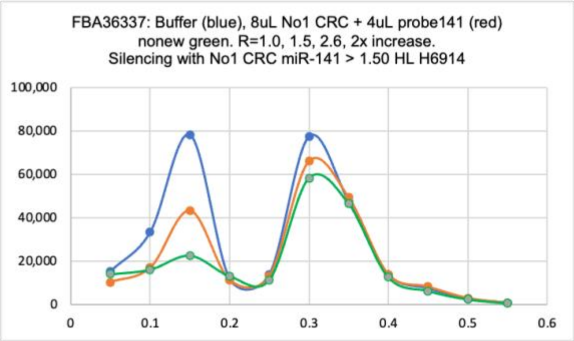
FBA36337: Buffer (blue), 8uL No1 CRC + 4uL probe141 (red)
nonew green. R=1.0, 1.5, 2.6, 2x increase.
Silencing with No1 CRC miR-141 > 1.50 HL H6914
100,000
80,000
60,000
40,000
20,000
0
0 0.1 0.2 0.3 0.4 0.5 0.6

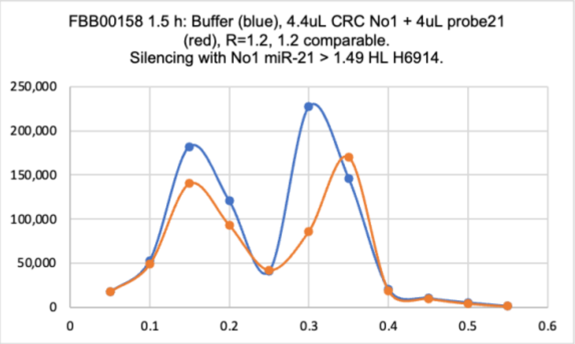
FBB00158 1.5 h: Buffer (blue), 4.4uL CRC No1 + 4uL probe21
(red), R=1.2, 1.2 comparable.
Silencing with No1 miR-21 > 1.49 HL H6914.
250,000
200,000
150,000
100,000
50,000
0
0 0.1 0.2 0.3 0.4 0.5 0.6

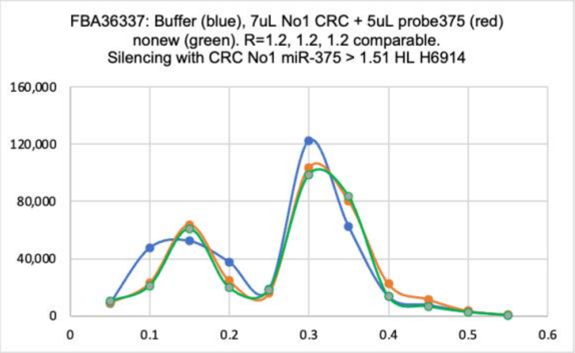
FBA36337: Buffer (blue), 7uL No1 CRC + 5uL probe375 (red)
nonew (green). R=1.2, 1.2, 1.2 comparable.
Silencing with CRC No1 miR-375 > 1.51 HL H6914
160,000
120,000
80,000
40,000
0
0 0.1 0.2 0.3 0.4 0.5 0.6

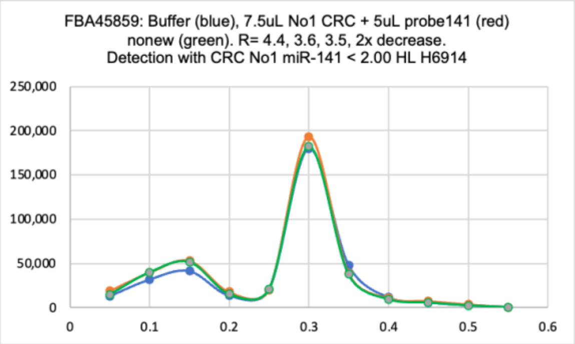
FBA45859: Buffer (blue), 7.5uL No1 CRC + 5uL probe141 (red)
nonew (green). R= 4.4, 3.6, 3.5, 2x decrease.
Detection with CRC No1 miR-141 < 2.00 HL H6914
250,000
200,000
150,000
100,000
50,000
0
0 0.1 0.2 0.3 0.4 0.5 0.6

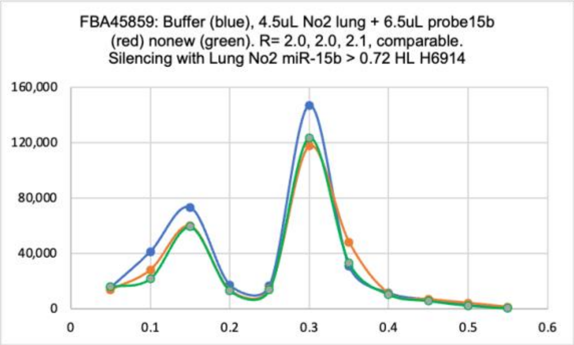

FBA45859: Buffer (blue), 4.5uL No2 lung + 6.5uL probe15b (red) nonew (green). R= 2.0, 2.0, 2.1, comparable. Silencing with Lung No2 miR-15b > 0.72 HL H6914

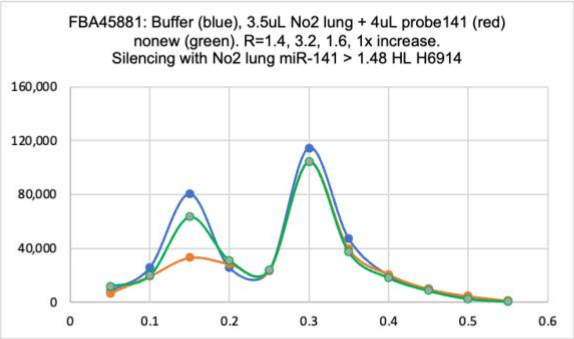

FBA45881: Buffer (blue), 3.5uL No2 lung + 4uL probe141 (red) nonew (green). R=1.4, 3.2, 1.6, 1x increase. Silencing with No2 lung miR-141 > 1.48 HL H6914

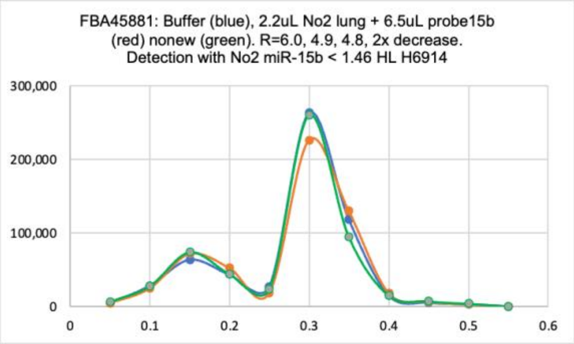

FBA45881: Buffer (blue), 2.2uL No2 lung + 6.5uL probe15b (red) nonew (green). R=6.0, 4.9, 4.8, 2x decrease. Detection with No2 miR-15b < 1.46 HL H6914

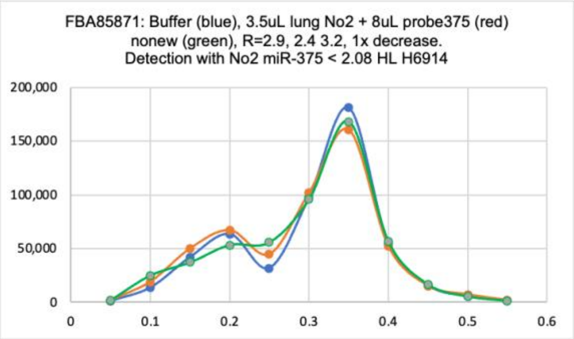

FBA85871: Buffer (blue), 3.5uL lung No2 + 8uL probe375 (red) nonew (green), R=2.9, 2.4 3.2, 1x decrease. Detection with No2 miR-375 < 2.08 HL H6914

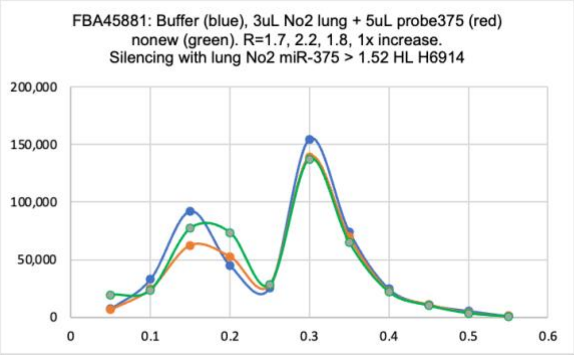

FBA45881: Buffer (blue), 3uL No2 lung + 5uL probe375 (red) nonew (green). R=1.7, 2.2, 1.8, 1x increase. Silencing with lung No2 miR-375 > 1.52 HL H6914

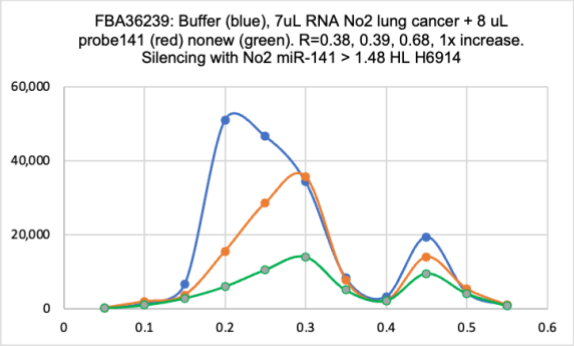

FBA36239: Buffer (blue), 7uL RNA No2 lung cancer + 8 uL probe141 (red) nonew (green). R=0.38, 0.39, 0.68, 1x increase. Silencing with No2 miR-141 > 1.48 HL H6914

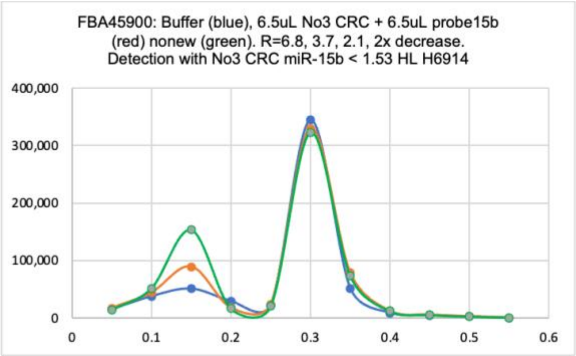
FBA45900: Buffer (blue), 6.5uL No3 CRC + 6.5uL probe15b
(red) nonew (green). R=6.8, 3.7, 2.1, 2x decrease.
Detection with No3 CRC miR-15b < 1.53 HL H6914
400,000
300,000
200,000
100,000
0
0 0.1 0.2 0.3 0.4 0.5 0.6

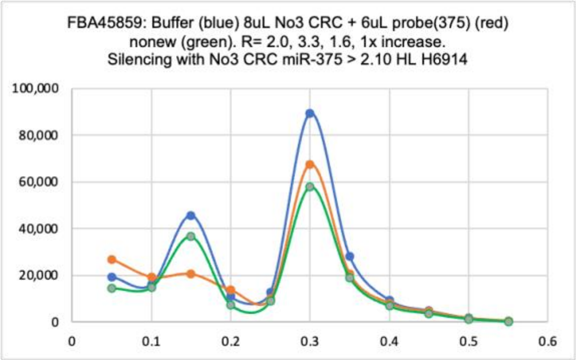
FBA45859: Buffer (blue) 8uL No3 CRC + 6uL probe(375) (red)
nonew (green). R= 2.0, 3.3, 1.6, 1x increase.
Silencing with No3 CRC miR-375 > 2.10 HL H6914
100,000
80,000
60,000
40,000
20,000
0
0 0.1 0.2 0.3 0.4 0.5 0.6

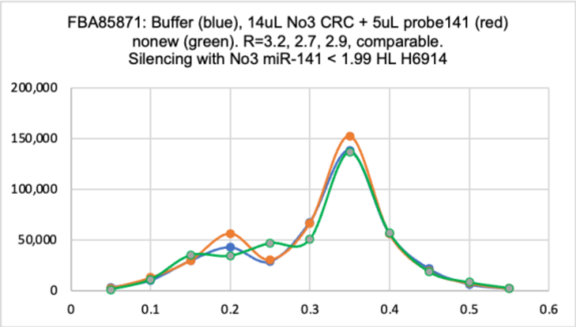
FBA85871: Buffer (blue), 14uL No3 CRC + 5uL probe141 (red)
nonew (green). R=3.2, 2.7, 2.9, comparable.
Silencing with No3 miR-141 < 1.99 HL H6914
200,000
150,000
100,000
50,000
0
0 0.1 0.2 0.3 0.4 0.5 0.6

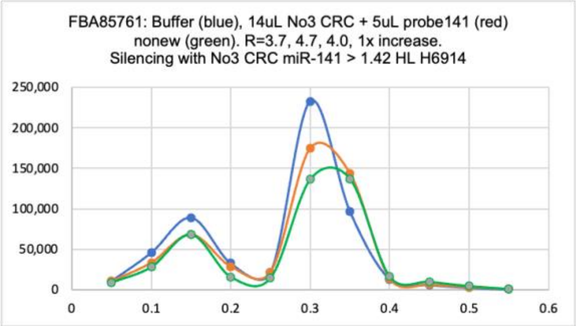
FBA85761: Buffer (blue), 14uL No3 CRC + 5uL probe141 (red)
nonew (green). R=3.7, 4.7, 4.0, 1x increase.
Silencing with No3 CRC miR-141 > 1.42 HL H6914
250,000
200,000
150,000
100,000
50,000
0
0 0.1 0.2 0.3 0.4 0.5 0.6

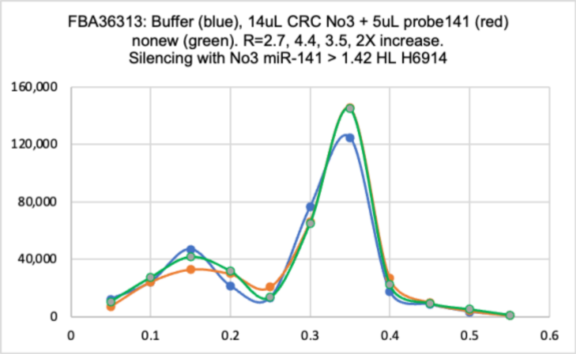
FBA36313: Buffer (blue), 14uL CRC No3 + 5uL probe141 (red)
nonew (green). R=2.7, 4.4, 3.5, 2X increase.
Silencing with No3 miR-141 > 1.42 HL H6914
160,000
120,000
80,000
40,000
0
0 0.1 0.2 0.3 0.4 0.5 0.6

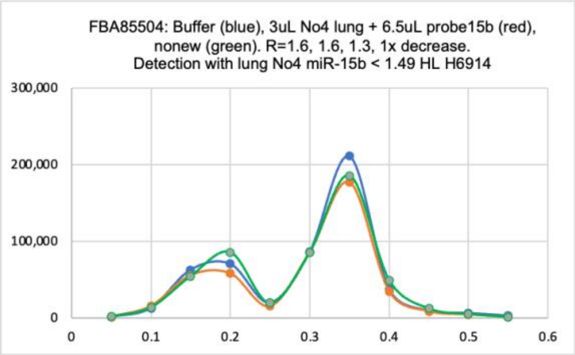
FBA85504: Buffer (blue), 3uL No4 lung + 6.5uL probe15b (red),
nonew (green). R=1.6, 1.6, 1.3, 1x decrease.
Detection with lung No4 miR-15b < 1.49 HL H6914
300,000
200,000
100,000
0
0 0.1 0.2 0.3 0.4 0.5 0.6

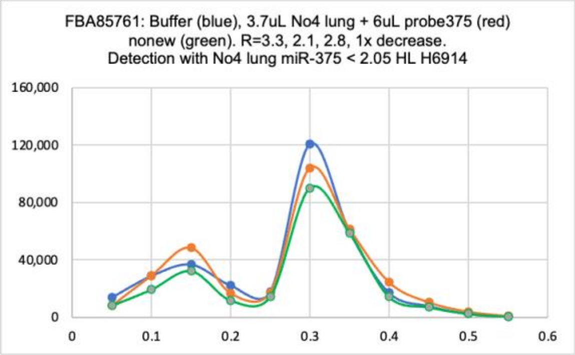
FBA85761: Buffer (blue), 3.7uL No4 lung + 6uL probe375 (red)
nonew (green). R=3.3, 2.1, 2.8, 1x decrease.
Detection with No4 lung miR-375 < 2.05 HL H6914
160,000
120,000
80,000
40,000
0
0 0.1 0.2 0.3 0.4 0.5 0.6

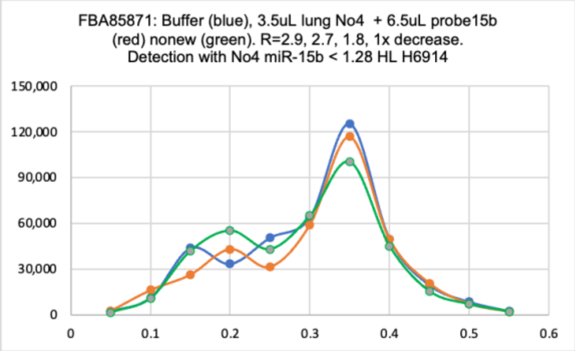
FBA85871: Buffer (blue), 3.5uL lung No4 + 6.5uL probe15b
(red) nonew (green). R=2.9, 2.7, 1.8, 1x decrease.
Detection with No4 miR-15b < 1.28 HL H6914
150,000
120,000
90,000
60,000
30,000
0
0 0.1 0.2 0.3 0.4 0.5 0.6

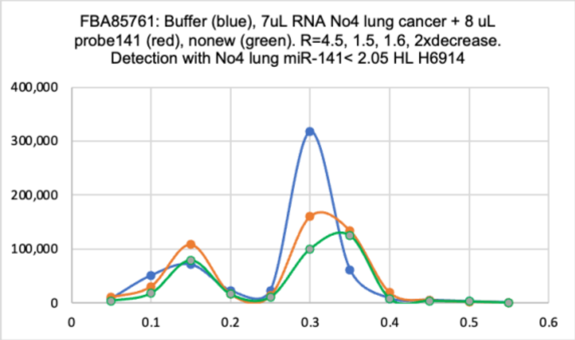
FBA85761: Buffer (blue), 7uL RNA No4 lung cancer + 8 uL
probe141 (red), nonew (green). R=4.5, 1.5, 1.6, 2xdecrease.
Detection with No4 lung miR-141< 2.05 HL H6914
400,000
300,000
200,000
100,000
0
0 0.1 0.2 0.3 0.4 0.5 0.6

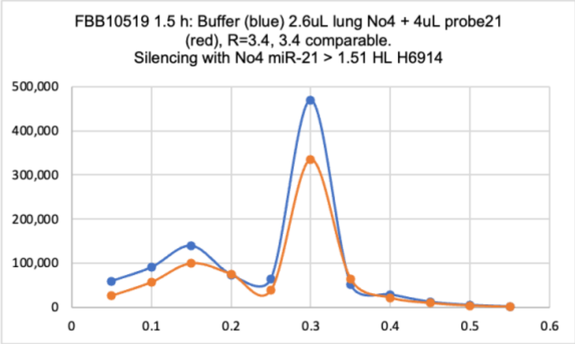
FBB10519 1.5 h: Buffer (blue) 2.6uL lung No4 + 4uL probe21
(red), R=3.4, 3.4 comparable.
Silencing with No4 miR-21 > 1.51 HL H6914
500,000
400,000
300,000
200,000
100,000
0
0 0.1 0.2 0.3 0.4 0.5 0.6

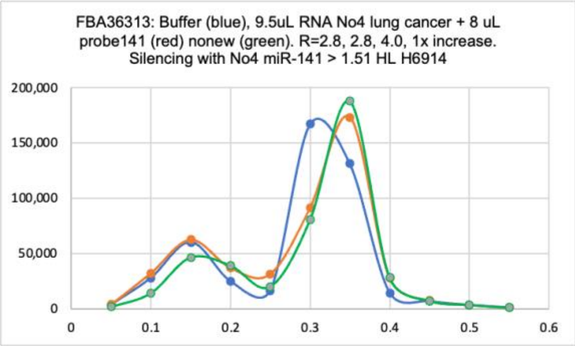
FBA36313: Buffer (blue), 9.5uL RNA No4 lung cancer + 8 uL
probe141 (red) nonew (green). R=2.8, 2.8, 4.0, 1x increase.
Silencing with No4 miR-141 > 1.51 HL H6914
200,000
150,000
100,000
50,000
0
0 0.1 0.2 0.3 0.4 0.5 0.6

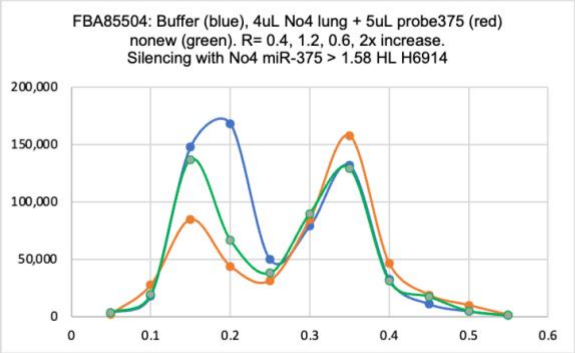
FBA85504: Buffer (blue), 4uL No4 lung + 5uL probe375 (red)
nonew (green). R= 0.4, 1.2, 0.6, 2x increase.
Silencing with No4 miR-375 > 1.58 HL H6914
200,000
150,000
100,000
50,000
0
0 0.1 0.2 0.3 0.4 0.5 0.6

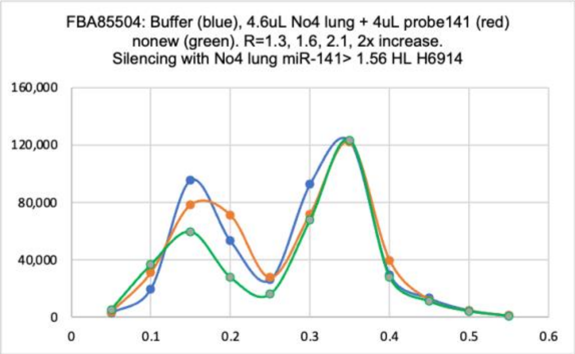
FBA85504: Buffer (blue), 4.6uL No4 lung + 4uL probe141 (red)
nonew (green). R=1.3, 1.6, 2.1, 2x increase.
Silencing with No4 lung miR-141> 1.56 HL H6914
160,000
120,000
80,000
40,000
0
0 0.1 0.2 0.3 0.4 0.5 0.6

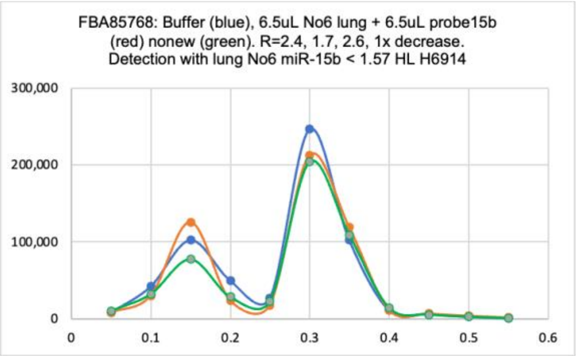
FBA85768: Buffer (blue), 6.5uL No6 lung + 6.5uL probe15b (red) nonew (green). R=2.4, 1.7, 2.6, 1x decrease.
Detection with lung No6 miR-15b < 1.57 HL H6914
300,000
200,000
100,000
0
0 0.1 0.2 0.3 0.4 0.5 0.6

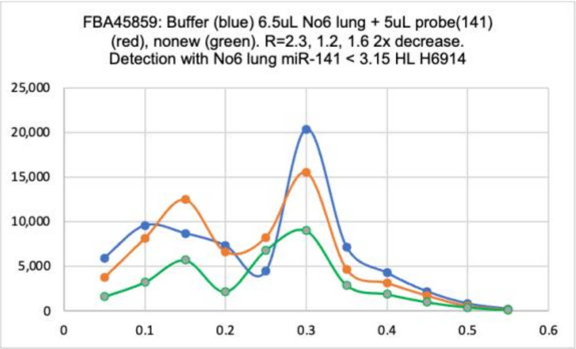
FBA45859: Buffer (blue) 6.5uL No6 lung + 5uL probe(141) (red), nonew (green). R=2.3, 1.2, 1.6 2x decrease.
Detection with No6 lung miR-141 < 3.15 HL H6914
25,000
20,000
15,000
10,000
5,000
0
0 0.1 0.2 0.3 0.4 0.5 0.6

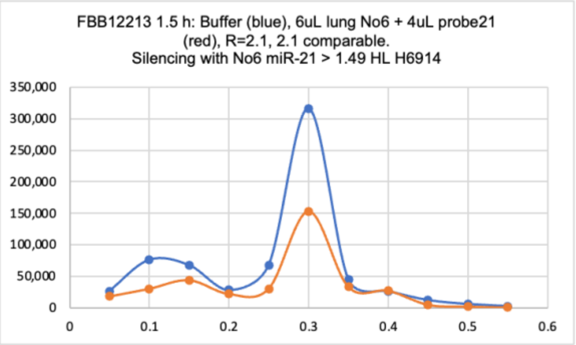
FBB12213 1.5 h: Buffer (blue), 6uL lung No6 + 4uL probe21 (red), R=2.1, 2.1 comparable.
Silencing with No6 miR-21 > 1.49 HL H6914
350,000
300,000
250,000
200,000
150,000
100,000
50,000
0
0 0.1 0.2 0.3 0.4 0.5 0.6

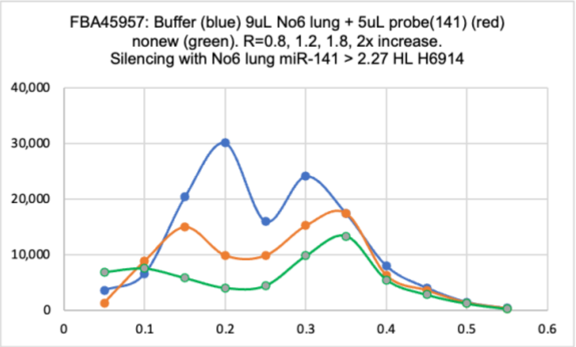
FBA45957: Buffer (blue) 9uL No6 lung + 5uL probe(141) (red) nonew (green). R=0.8, 1.2, 1.8, 2x increase.
Silencing with No6 lung miR-141 > 2.27 HL H6914
40,000
30,000
20,000
10,000
0
0 0.1 0.2 0.3 0.4 0.5 0.6

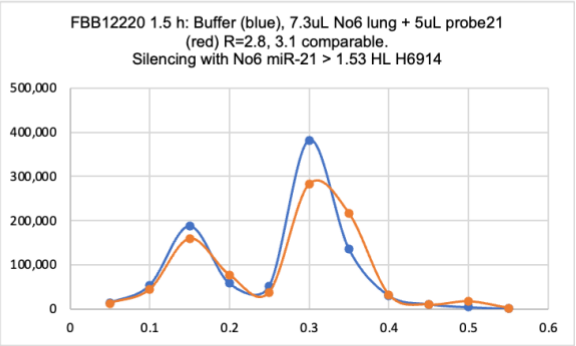
FBB12220 1.5 h: Buffer (blue), 7.3uL No6 lung + 5uL probe21 (red) R=2.8, 3.1 comparable.
Silencing with No6 miR-21 > 1.53 HL H6914
500,000
400,000
300,000
200,000
100,000
0
0 0.1 0.2 0.3 0.4 0.5 0.6

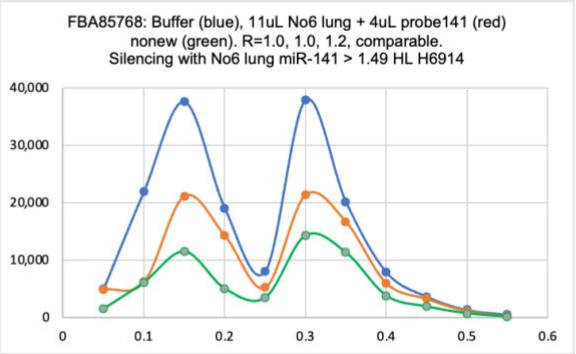
FBA85768: Buffer (blue), 11uL No6 lung + 4uL probe141 (red) nonew (green). R=1.0, 1.0, 1.2, comparable.
Silencing with No6 lung miR-141 > 1.49 HL H6914
40,000
30,000
20,000
10,000
0
0 0.1 0.2 0.3 0.4 0.5 0.6

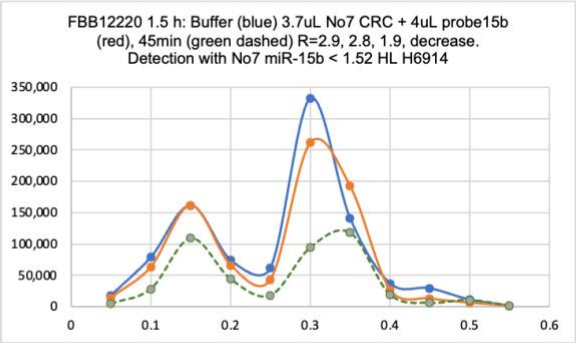
FBB12220 1.5 h: Buffer (blue) 3.7uL No7 CRC + 4uL probe15b (red), 45min (green dashed) R=2.9, 2.8, 1.9, decrease.
Detection with No7 miR-15b < 1.52 HL H6914
350,000
300,000
250,000
200,000
150,000
100,000
50,000
0
0 0.1 0.2 0.3 0.4 0.5 0.6

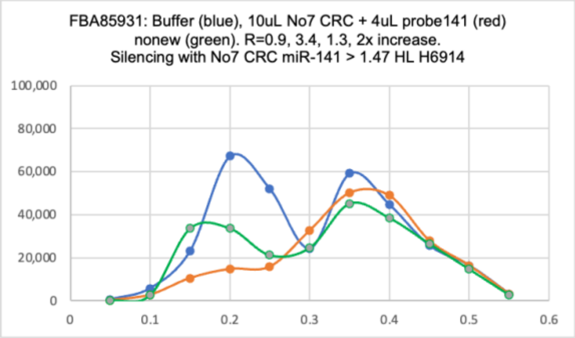
FBA85931: Buffer (blue), 10uL No7 CRC + 4uL probe141 (red) nonew (green). R=0.9, 3.4, 1.3, 2x increase.
Silencing with No7 CRC miR-141 > 1.47 HL H6914
100,000
80,000
60,000
40,000
20,000
0
0 0.1 0.2 0.3 0.4 0.5 0.6

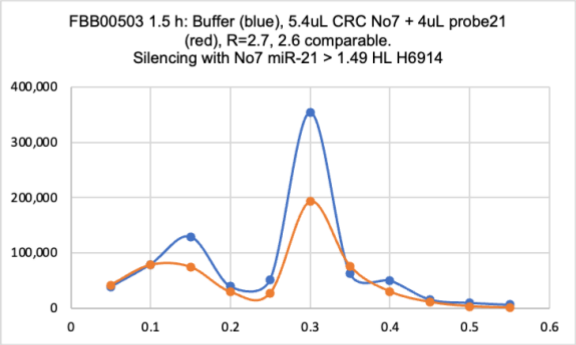
FBB00503 1.5 h: Buffer (blue), 5.4uL CRC No7 + 4uL probe21 (red), R=2.7, 2.6 comparable.
Silencing with No7 miR-21 > 1.49 HL H6914
400,000
300,000
200,000
100,000
0
0 0.1 0.2 0.3 0.4 0.5 0.6

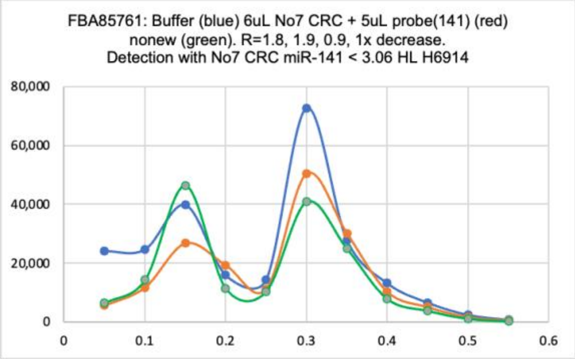
FBA85761: Buffer (blue) 6uL No7 CRC + 5uL probe(141) (red) nonew (green). R=1.8, 1.9, 0.9, 1x decrease.
Detection with No7 CRC miR-141 < 3.06 HL H6914
80,000
60,000
40,000
20,000
0
0 0.1 0.2 0.3 0.4 0.5 0.6

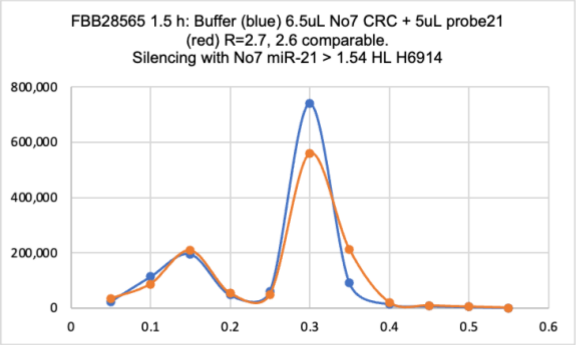
FBB28565 1.5 h: Buffer (blue) 6.5uL No7 CRC + 5uL probe21 (red) R=2.7, 2.6 comparable.
Silencing with No7 miR-21 > 1.54 HL H6914
800,000
600,000
400,000
200,000
0
0 0.1 0.2 0.3 0.4 0.5 0.6

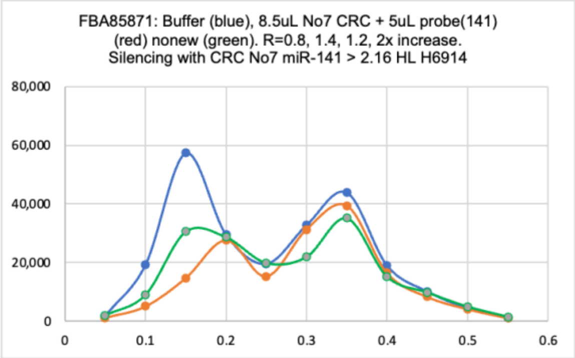
FBA85871: Buffer (blue), 8.5uL No7 CRC + 5uL probe(141) (red) nonew (green). R=0.8, 1.4, 1.2, 2x increase.
Silencing with CRC No7 miR-141 > 2.16 HL H6914
80,000
60,000
40,000
20,000
0
0 0.1 0.2 0.3 0.4 0.5 0.6

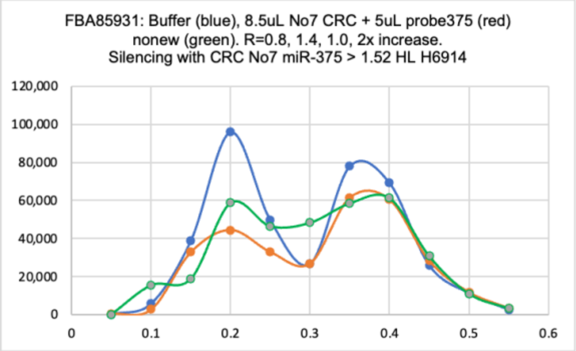
FBA85931: Buffer (blue), 8.5uL No7 CRC + 5uL probe375 (red) nonew (green). R=0.8, 1.4, 1.0, 2x increase.
Silencing with CRC No7 miR-375 > 1.52 HL H6914
120,000
100,000
80,000
60,000
40,000
20,000
0
0 0.1 0.2 0.3 0.4 0.5 0.6

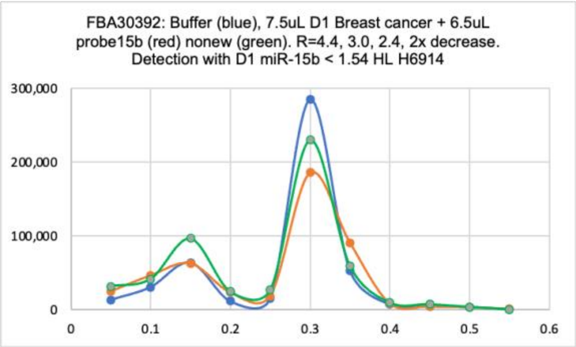
FBA30392: Buffer (blue), 7.5uL D1 Breast cancer + 6.5uL probe15b (red) nonew (green). R=4.4, 3.0, 2.4, 2x decrease. Detection with D1 miR-15b < 1.54 HL H6914
300,000
200,000
100,000
0
0 0.1 0.2 0.3 0.4 0.5 0.6

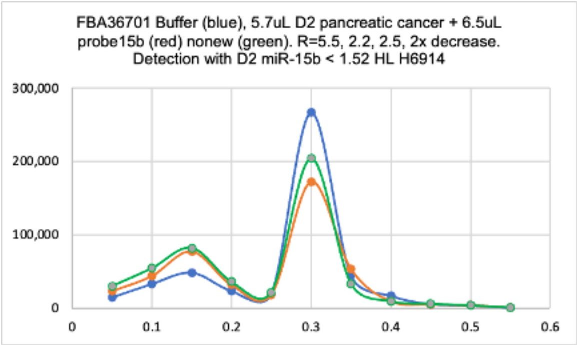
FBA36701 Buffer (blue), 5.7uL D2 pancreatic cancer + 6.5uL probe15b (red) nonew (green). R=5.5, 2.2, 2.5, 2x decrease. Detection with D2 miR-15b < 1.52 HL H6914
300,000
200,000
100,000
0
0 0.1 0.2 0.3 0.4 0.5 0.6

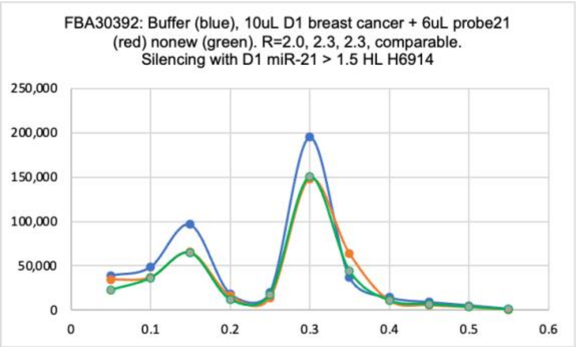
FBA30392: Buffer (blue), 10uL D1 breast cancer + 6uL probe21 (red) nonew (green). R=2.0, 2.3, 2.3, comparable. Silencing with D1 miR-21 > 1.5 HL H6914
250,000
200,000
150,000
100,000
50,000
0
0 0.1 0.2 0.3 0.4 0.5 0.6

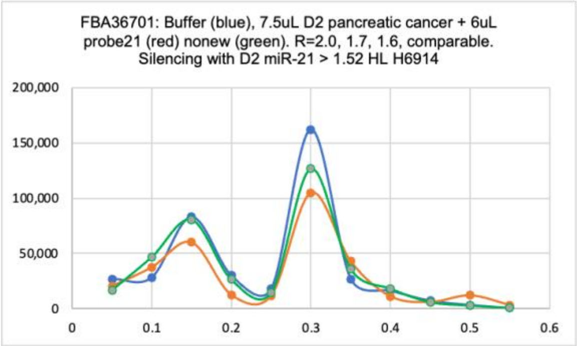
FBA36701: Buffer (blue), 7.5uL D2 pancreatic cancer + 6uL probe21 (red) nonew (green). R=2.0, 1.7, 1.6, comparable. Silencing with D2 miR-21 > 1.52 HL H6914
200,000
150,000
100,000
50,000
0
0 0.1 0.2 0.3 0.4 0.5 0.6

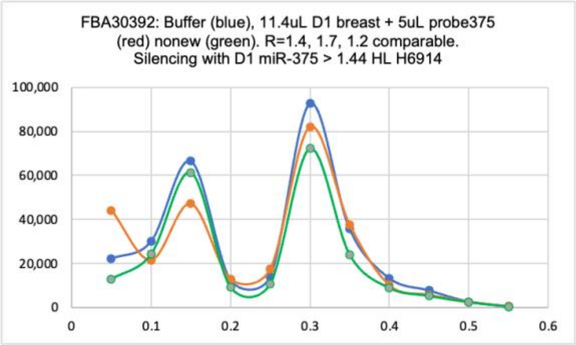
FBA30392: Buffer (blue), 11.4uL D1 breast + 5uL probe375 (red) nonew (green). R=1.4, 1.7, 1.2 comparable. Silencing with D1 miR-375 > 1.44 HL H6914
100,000
80,000
60,000
40,000
20,000
0
0 0.1 0.2 0.3 0.4 0.5 0.6

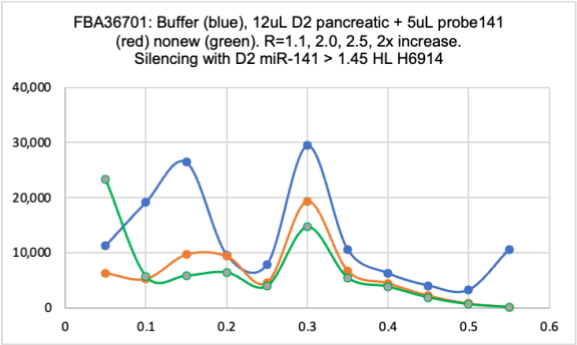
FBA36701: Buffer (blue), 12uL D2 pancreatic + 5uL probe141 (red) nonew (green). R=1.1, 2.0, 2.5, 2x increase. Silencing with D2 miR-141 > 1.45 HL H6914
40,000
30,000
20,000
10,000
0
0 0.1 0.2 0.3 0.4 0.5 0.6

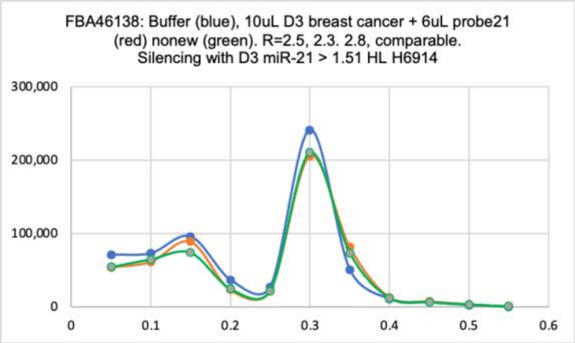


FBA46138: Buffer (blue), 10uL D3 breast cancer + 6uL probe21 (red) nonew (green). R=2.5, 2.3. 2.8, comparable.
Silencing with D3 miR-21 > 1.51 HL H6914

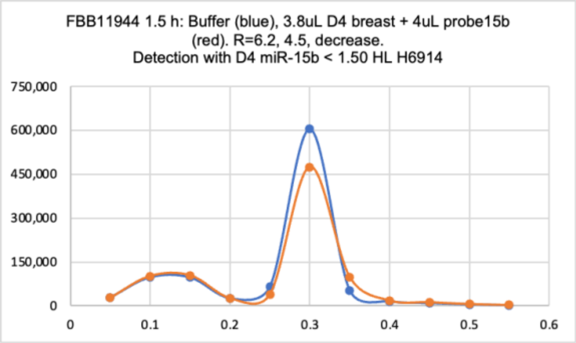


FBB11944 1.5 h: Buffer (blue), 3.8uL D4 breast + 4uL probe15b (red). R=6.2, 4.5, decrease.
Detection with D4 miR-15b < 1.50 HL H6914

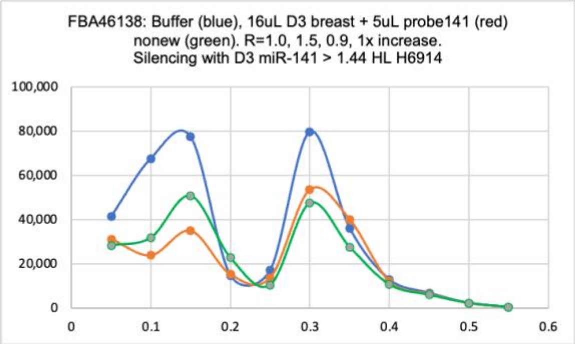


FBA46138: Buffer (blue), 16uL D3 breast + 5uL probe141 (red) nonew (green). R=1.0, 1.5, 0.9, 1x increase.
Silencing with D3 miR-141 > 1.44 HL H6914

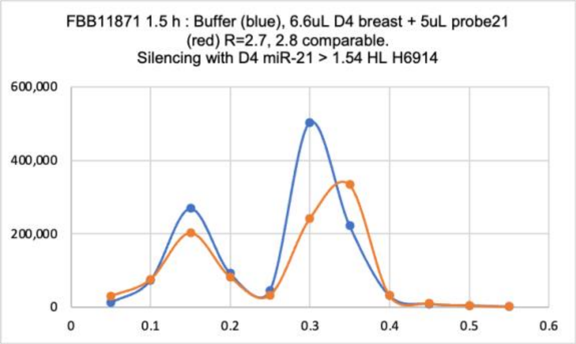


FBB11871 1.5 h : Buffer (blue), 6.6uL D4 breast + 5uL probe21 (red) R=2.7, 2.8 comparable.
Silencing with D4 miR-21 > 1.54 HL H6914

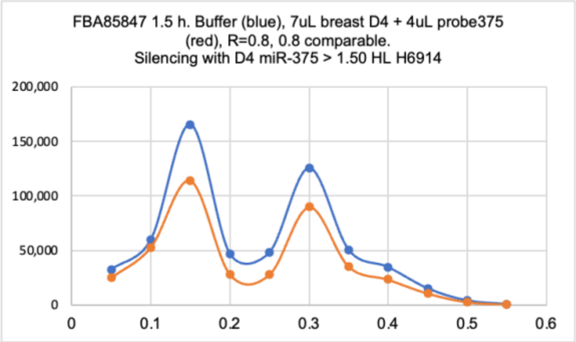


FBA85847 1.5 h. Buffer (blue), 7uL breast D4 + 4uL probe375 (red), R=0.8, 0.8 comparable.
Silencing with D4 miR-375 > 1.50 HL H6914

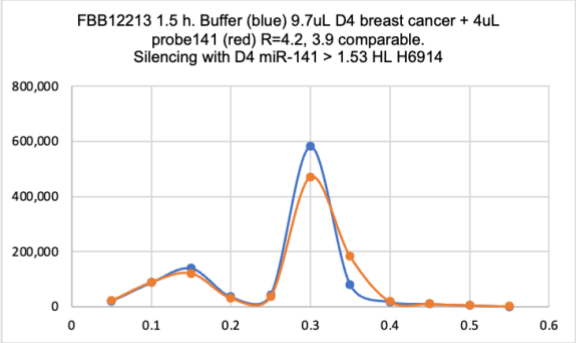


FBB12213 1.5 h. Buffer (blue) 9.7uL D4 breast cancer + 4uL probe141 (red) R=4.2, 3.9 comparable.
Silencing with D4 miR-141 > 1.53 HL H6914

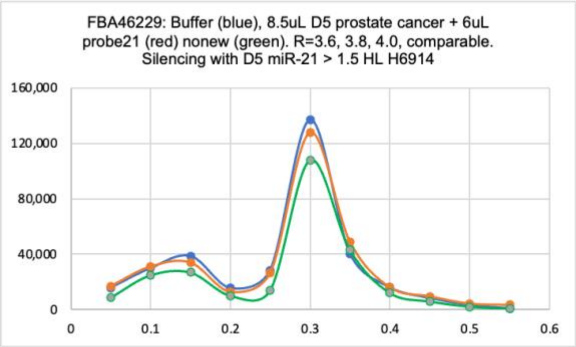
FBA46229: Buffer (blue), 8.5uL D5 prostate cancer + 6uL probe21 (red) nonew (green). R=3.6, 3.8, 4.0, comparable.
Silencing with D5 miR-21 > 1.5 HL H6914
160,000
120,000
80,000
40,000
0
0 0.1 0.2 0.3 0.4 0.5 0.6

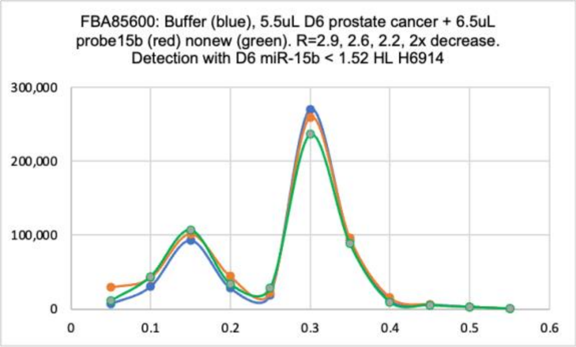
FBA85600: Buffer (blue), 5.5uL D6 prostate cancer + 6.5uL probe15b (red) nonew (green). R=2.9, 2.6, 2.2, 2x decrease.
Detection with D6 miR-15b < 1.52 HL H6914
300,000
200,000
100,000
0
0 0.1 0.2 0.3 0.4 0.5 0.6

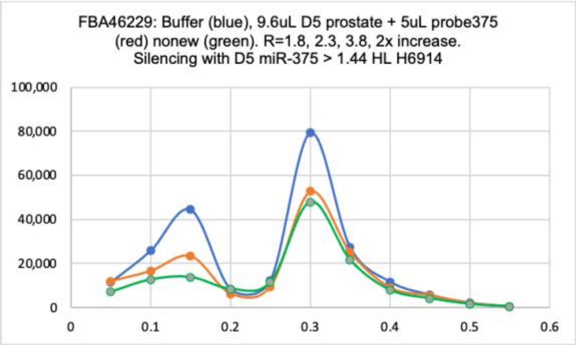
FBA46229: Buffer (blue), 9.6uL D5 prostate + 5uL probe375 (red) nonew (green). R=1.8, 2.3, 3.8, 2x increase.
Silencing with D5 miR-375 > 1.44 HL H6914
100,000
80,000
60,000
40,000
20,000
0
0 0.1 0.2 0.3 0.4 0.5 0.6

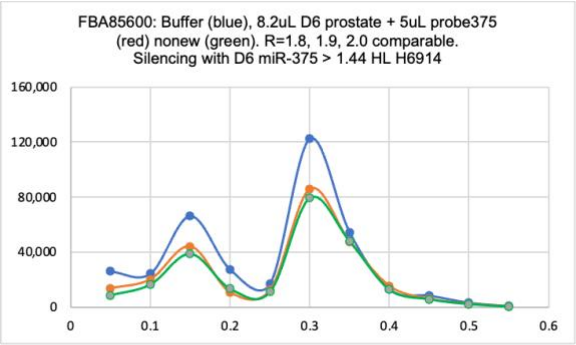
FBA85600: Buffer (blue), 8.2uL D6 prostate + 5uL probe375 (red) nonew (green). R=1.8, 1.9, 2.0 comparable.
Silencing with D6 miR-375 > 1.44 HL H6914
160,000
120,000
80,000
40,000
0
0 0.1 0.2 0.3 0.4 0.5 0.6

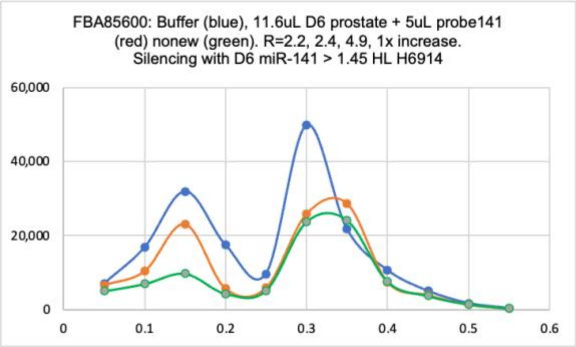
FBA85600: Buffer (blue), 11.6uL D6 prostate + 5uL probe141 (red) nonew (green). R=2.2, 2.4, 4.9, 1x increase.
Silencing with D6 miR-141 > 1.45 HL H6914
60,000
40,000
20,000
0
0 0.1 0.2 0.3 0.4 0.5 0.6

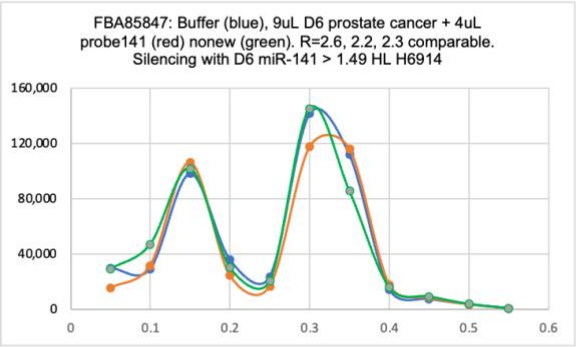
FBA85847: Buffer (blue), 9uL D6 prostate cancer + 4uL probe141 (red) nonew (green). R=2.6, 2.2, 2.3 comparable.
Silencing with D6 miR-141 > 1.49 HL H6914
160,000
120,000
80,000
40,000
0
0 0.1 0.2 0.3 0.4 0.5 0.6

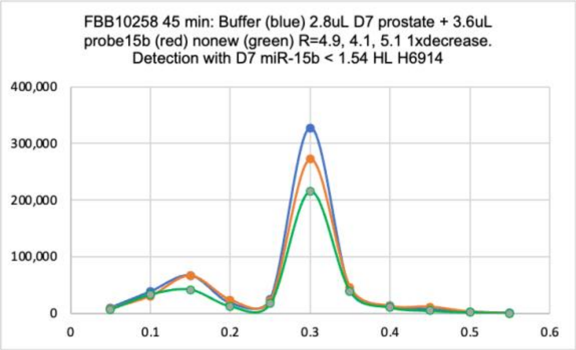

FBB10258 45 min: Buffer (blue) 2.8uL D7 prostate + 3.6uL probe15b (red) nonew (green) R=4.9, 4.1, 5.1 1xdecrease. Detection with D7 miR-15b < 1.54 HL H6914

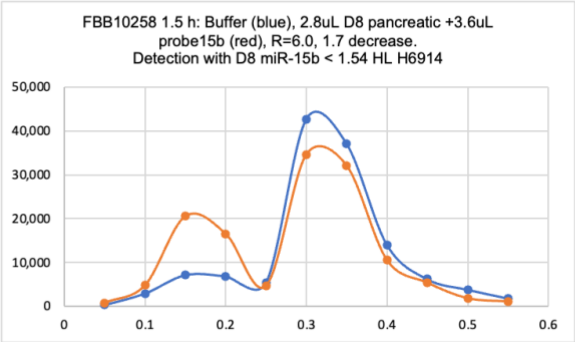

FBB10258 1.5 h: Buffer (blue), 2.8uL D8 pancreatic +3.6uL probe15b (red), R=6.0, 1.7 decrease. Detection with D8 miR-15b < 1.54 HL H6914

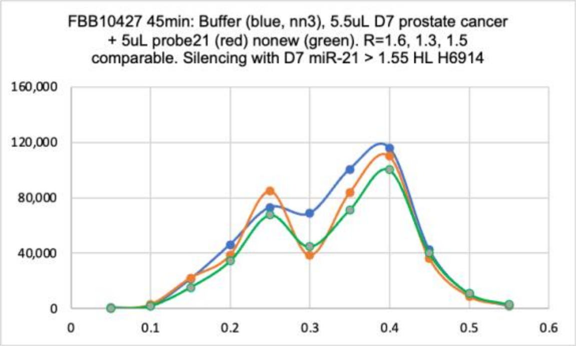

FBB10427 45min: Buffer (blue, nn3), 5.5uL D7 prostate cancer + 5uL probe21 (red) nonew (green). R=1.6, 1.3, 1.5 comparable. Silencing with D7 miR-21 > 1.55 HL H6914

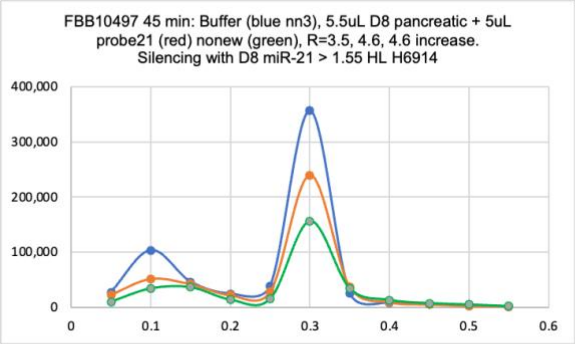

FBB10497 45 min: Buffer (blue nn3), 5.5uL D8 pancreatic + 5uL probe21 (red) nonew (green), R=3.5, 4.6, 4.6 increase. Silencing with D8 miR-21 > 1.55 HL H6914

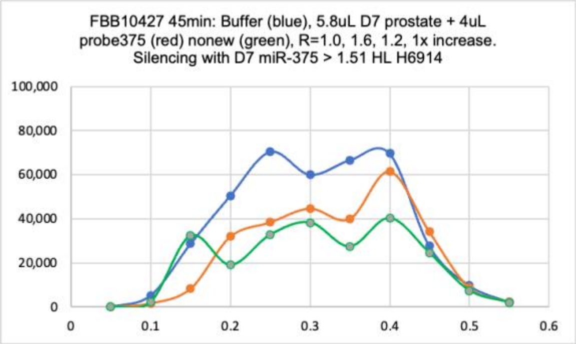

FBB10427 45min: Buffer (blue), 5.8uL D7 prostate + 4uL probe375 (red) nonew (green), R=1.0, 1.6, 1.2, 1x increase. Silencing with D7 miR-375 > 1.51 HL H6914

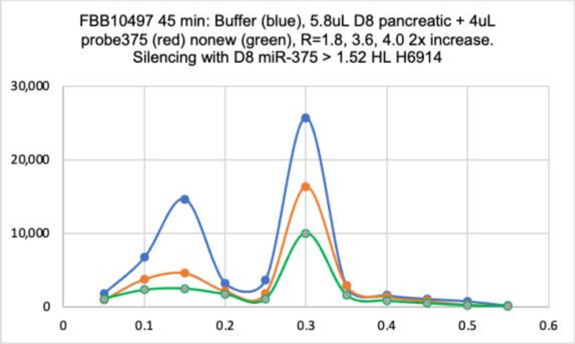

FBB10497 45 min: Buffer (blue), 5.8uL D8 pancreatic + 4uL probe375 (red) nonew (green), R=1.8, 3.6, 4.0 2x increase. Silencing with D8 miR-375 > 1.52 HL H6914

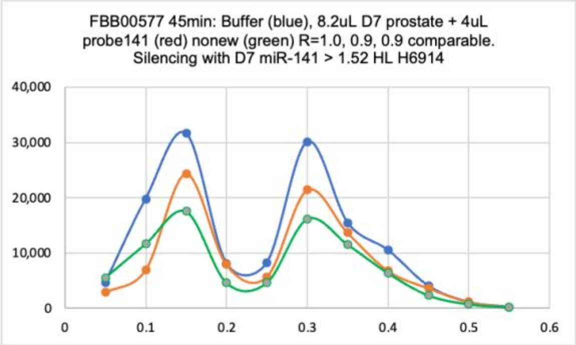

FBB00577 45min: Buffer (blue), 8.2uL D7 prostate + 4uL probe141 (red) nonew (green) R=1.0, 0.9, 0.9 comparable. Silencing with D7 miR-141 > 1.52 HL H6914

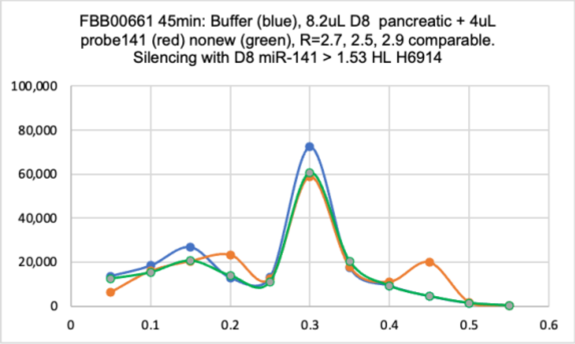

FBB00661 45min: Buffer (blue), 8.2uL D8 pancreatic + 4uL probe141 (red) nonew (green), R=2.7, 2.5, 2.9 comparable. Silencing with D8 miR-141 > 1.53 HL H6914

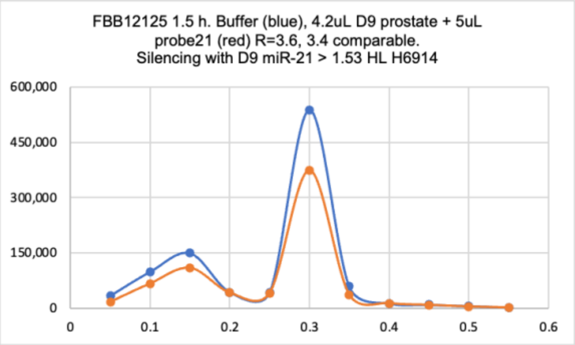
FBB12125 1.5 h. Buffer (blue), 4.2uL D9 prostate + 5uL
probe21 (red) R=3.6, 3.4 comparable.
Silencing with D9 miR-21 > 1.53 HL H6914
600,000
450,000
300,000
150,000
0
0 0.1 0.2 0.3 0.4 0.5 0.6

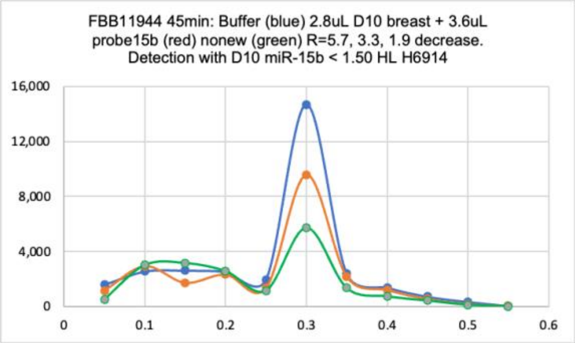
FBB11944 45min: Buffer (blue) 2.8uL D10 breast + 3.6uL
probe15b (red) nonew (green) R=5.7, 3.3, 1.9 decrease.
Detection with D10 miR-15b < 1.50 HL H6914
16,000
12,000
8,000
4,000
0
0 0.1 0.2 0.3 0.4 0.5 0.6

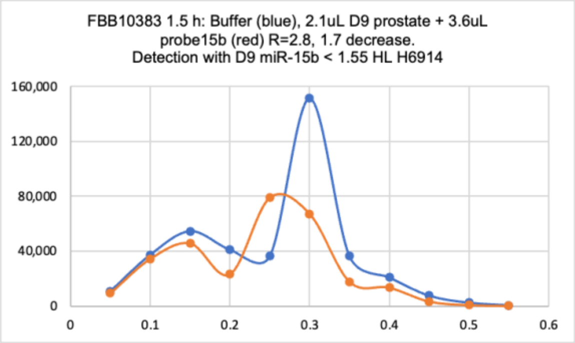
FBB10383 1.5 h: Buffer (blue), 2.1uL D9 prostate + 3.6uL
probe15b (red) R=2.8, 1.7 decrease.
Detection with D9 miR-15b < 1.55 HL H6914
160,000
120,000
80,000
40,000
0
0 0.1 0.2 0.3 0.4 0.5 0.6

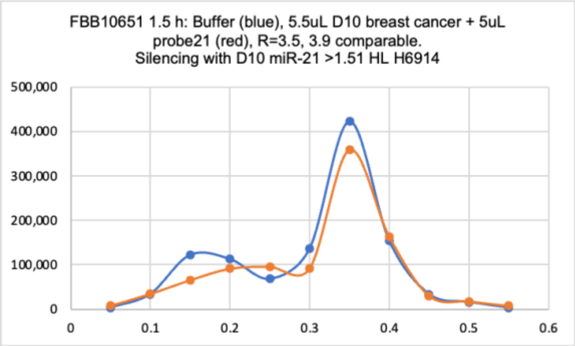
FBB10651 1.5 h: Buffer (blue), 5.5uL D10 breast cancer + 5uL
probe21 (red), R=3.5, 3.9 comparable.
Silencing with D10 miR-21 >1.51 HL H6914
500,000
400,000
300,000
200,000
100,000
0
0 0.1 0.2 0.3 0.4 0.5 0.6

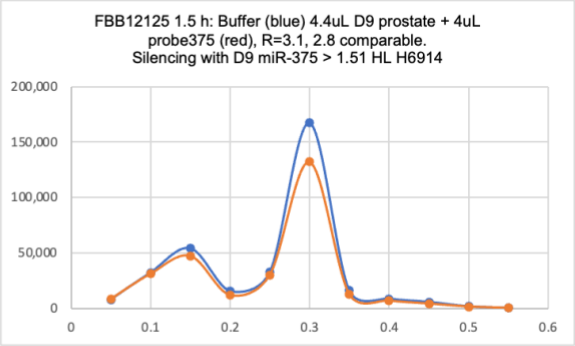
FBB12125 1.5 h: Buffer (blue) 4.4uL D9 prostate + 4uL
probe375 (red), R=3.1, 2.8 comparable.
Silencing with D9 miR-375 > 1.51 HL H6914
200,000
150,000
100,000
50,000
0
0 0.1 0.2 0.3 0.4 0.5 0.6

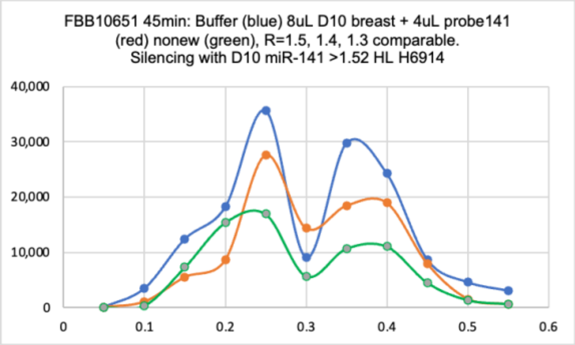
FBB10651 45min: Buffer (blue) 8uL D10 breast + 4uL probe141
(red) nonew (green), R=1.5, 1.4, 1.3 comparable.
Silencing with D10 miR-141 >1.52 HL H6914
40,000
30,000
20,000
10,000
0
0 0.1 0.2 0.3 0.4 0.5 0.6

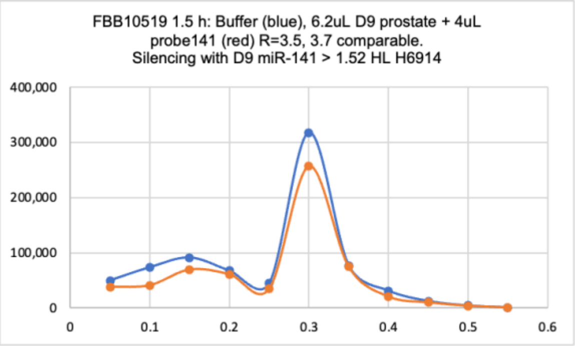
FBB10519 1.5 h: Buffer (blue), 6.2uL D9 prostate + 4uL
probe141 (red) R=3.5, 3.7 comparable.
Silencing with D9 miR-141 > 1.52 HL H6914
400,000
300,000
200,000
100,000
0
0 0.1 0.2 0.3 0.4 0.5 0.6

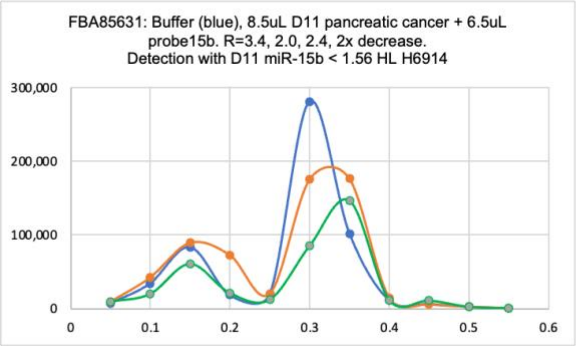
FBA85631: Buffer (blue), 8.5uL D11 pancreatic cancer + 6.5uL probe15b. R=3.4, 2.0, 2.4, 2x decrease.
Detection with D11 miR-15b < 1.56 HL H6914
300,000
200,000
100,000
0
0
0.1
0.2
0.3
0.4
0.5
0.6

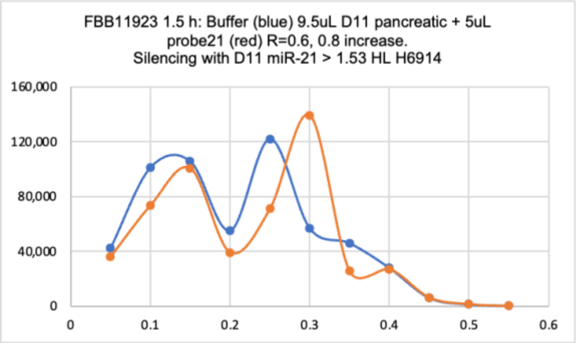
FBB11923 1.5 h: Buffer (blue) 9.5uL D11 pancreatic + 5uL probe21 (red) R=0.6, 0.8 increase.
Silencing with D11 miR-21 > 1.53 HL H6914
160,000
120,000
80,000
40,000
0
0
0.1
0.2
0.3
0.4
0.5
0.6

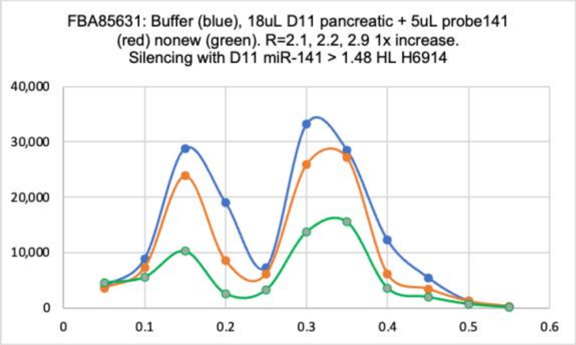
FBA85631: Buffer (blue), 18uL D11 pancreatic + 5uL probe141 (red) nonew (green). R=2.1, 2.2, 2.9 1x increase.
Silencing with D11 miR-141 > 1.48 HL H6914
40,000
30,000
20,000
10,000
0
0
0.1
0.2
0.3
0.4
0.5
0.6

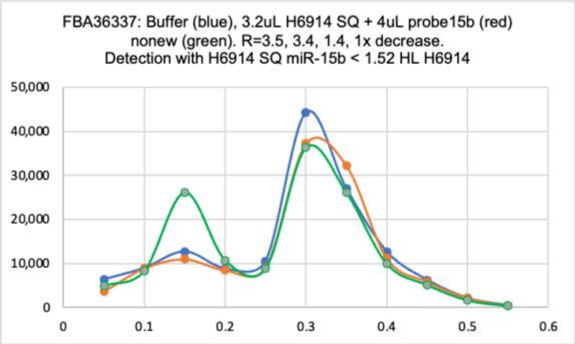
FBA36337: Buffer (blue), 3.2uL H6914 SQ + 4uL probe15b (red)
nonew (green). R=3.5, 3.4, 1.4, 1x decrease.
Detection with H6914 SQ miR-15b < 1.52 HL H6914
50,000 40,000 30,000 20,000 10,000 0
0 0.1 0.2 0.3 0.4 0.5 0.6

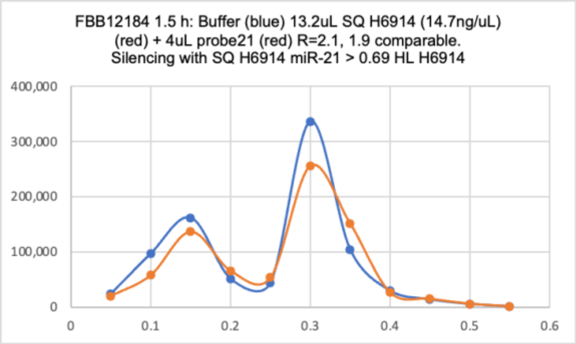
FBB12184 1.5 h: Buffer (blue) 13.2uL SQ H6914 (14.7ng/uL)
(red) + 4uL probe21 (red) R=2.1, 1.9 comparable.
Silencing with SQ H6914 miR-21 > 0.69 HL H6914
400,000 300,000 200,000 100,000 0
0 0.1 0.2 0.3 0.4 0.5 0.6

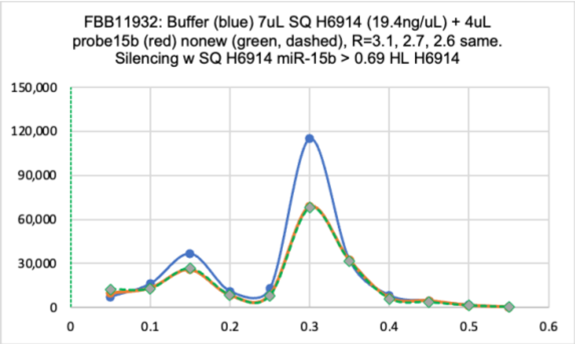
FBB11932: Buffer (blue) 7uL SQ H6914 (19.4ng/uL) + 4uL
probe15b (red) nonew (green, dashed), R=3.1, 2.7, 2.6 same.
Silencing w SQ H6914 miR-15b > 0.69 HL H6914
150,000 120,000 90,000 60,000 30,000 0
0 0.1 0.2 0.3 0.4 0.5 0.6

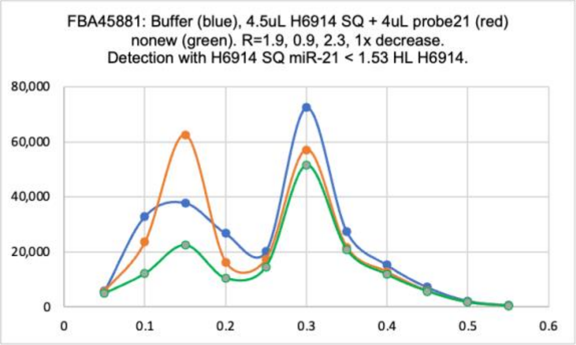
FBA45881: Buffer (blue), 4.5uL H6914 SQ + 4uL probe21 (red)
nonew (green). R=1.9, 0.9, 2.3, 1x decrease.
Detection with H6914 SQ miR-21 < 1.53 HL H6914.
80,000 60,000 40,000 20,000 0
0 0.1 0.2 0.3 0.4 0.5 0.6

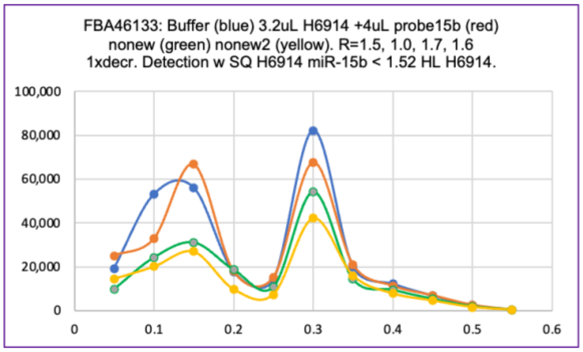
FBA46133: Buffer (blue) 3.2uL H6914 +4uL probe15b (red)
nonew (green) nonew2 (yellow). R=1.5, 1.0, 1.7, 1.6
1xdecr. Detection w SQ H6914 miR-15b < 1.52 HL H6914.
100,000 80,000 60,000 40,000 20,000 0
0 0.1 0.2 0.3 0.4 0.5 0.6

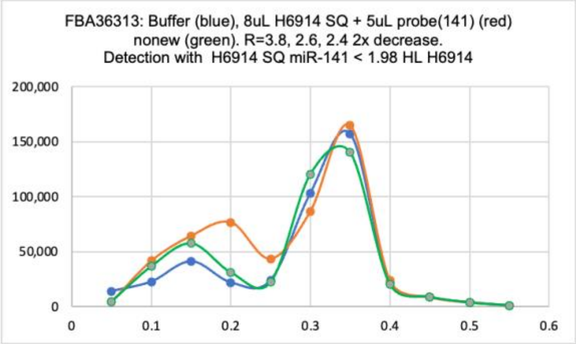
FBA36313: Buffer (blue), 8uL H6914 SQ + 5uL probe(141) (red)
nonew (green). R=3.8, 2.6, 2.4 2x decrease.
Detection with H6914 SQ miR-141 < 1.98 HL H6914
200,000 150,000 100,000 50,000 0
0 0.1 0.2 0.3 0.4 0.5 0.6

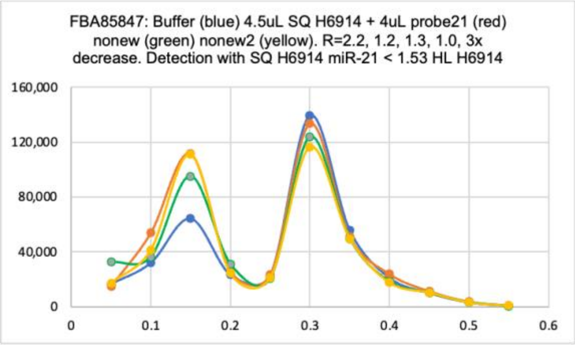
FBA85847: Buffer (blue) 4.5uL SQ H6914 + 4uL probe21 (red)
nonew (green) nonew2 (yellow). R=2.2, 1.2, 1.3, 1.0, 3x
decrease. Detection with SQ H6914 miR-21 < 1.53 HL H6914
160,000 120,000 80,000 40,000 0
0 0.1 0.2 0.3 0.4 0.5 0.6

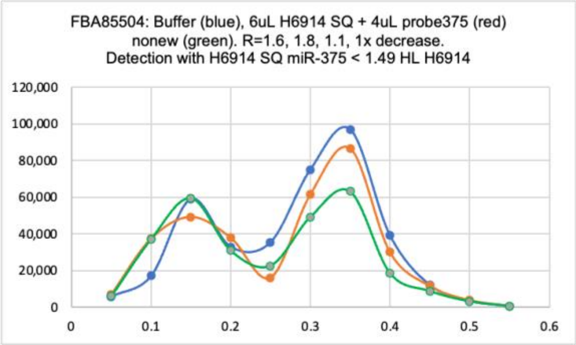
FBA85504: Buffer (blue), 6uL H6914 SQ + 4uL probe375 (red)
nonew (green). R=1.6, 1.8, 1.1, 1x decrease.
Detection with H6914 SQ miR-375 < 1.49 HL H6914
120,000 100,000 80,000 60,000 40,000 20,000 0
0 0.1 0.2 0.3 0.4 0.5 0.6

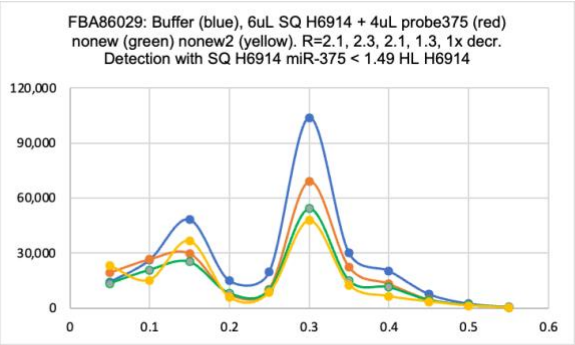
FBA86029: Buffer (blue), 6uL SQ H6914 + 4uL probe375 (red)
nonew (green) nonew2 (yellow). R=2.1, 2.3, 2.1, 1.3, 1x decr.
Detection with SQ H6914 miR-375 < 1.49 HL H6914
120,000
90,000
60,000
30,000
0
0 0.1 0.2 0.3 0.4 0.5 0.6

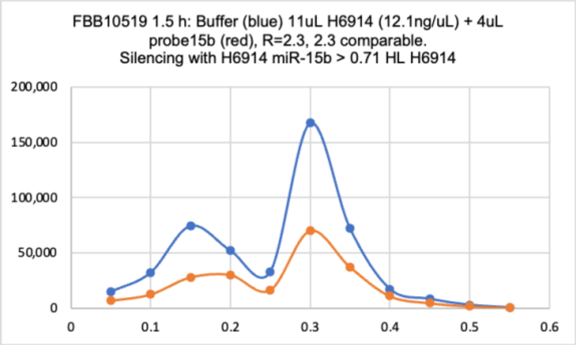
FBB10519 1.5 h: Buffer (blue) 11uL H6914 (12.1ng/uL) + 4uL
probe15b (red), R=2.3, 2.3 comparable.
Silencing with H6914 miR-15b > 0.71 HL H6914
200,000
150,000
100,000
50,000
0
0 0.1 0.2 0.3 0.4 0.5 0.6

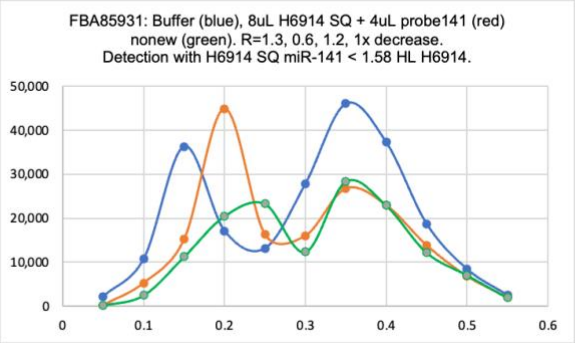
FBA85931: Buffer (blue), 8uL H6914 SQ + 4uL probe141 (red)
nonew (green). R=1.3, 0.6, 1.2, 1x decrease.
Detection with H6914 SQ miR-141 < 1.58 HL H6914.
50,000
40,000
30,000
20,000
10,000
0
0 0.1 0.2 0.3 0.4 0.5 0.6

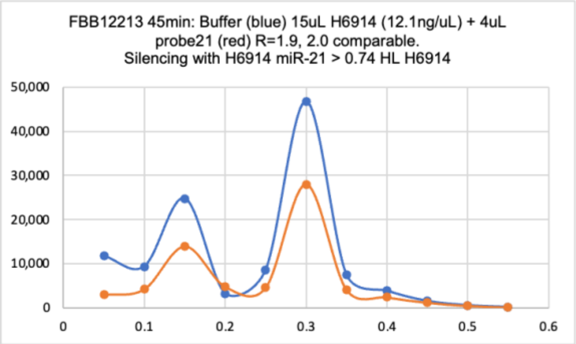
FBB12213 45min: Buffer (blue) 15uL H6914 (12.1ng/uL) + 4uL
probe21 (red) R=1.9, 2.0 comparable.
Silencing with H6914 miR-21 > 0.74 HL H6914
50,000
40,000
30,000
20,000
10,000
0
0 0.1 0.2 0.3 0.4 0.5 0.6

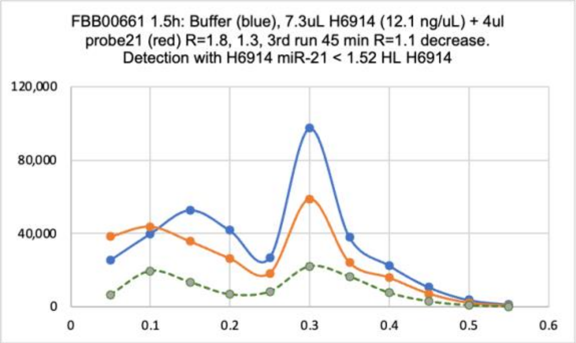
FBB00661 1.5h: Buffer (blue), 7.3uL H6914 (12.1 ng/uL) + 4ul
probe21 (red) R=1.8, 1.3, 3rd run 45 min R=1.1 decrease.
Detection with H6914 miR-21 < 1.52 HL H6914
120,000
80,000
40,000
0
0 0.1 0.2 0.3 0.4 0.5 0.6

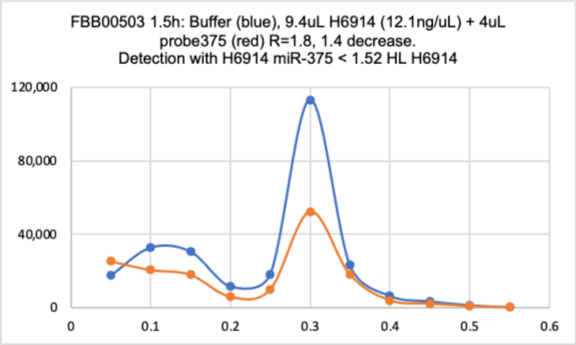
FBB00503 1.5h: Buffer (blue), 9.4uL H6914 (12.1ng/uL) + 4uL
probe375 (red) R=1.8, 1.4 decrease.
Detection with H6914 miR-375 < 1.52 HL H6914
120,000
80,000
40,000
0
0 0.1 0.2 0.3 0.4 0.5 0.6

FBB12184 45 min: Buffer (blue), 2.5uL Healthy1 urine + 5uL probe21 (red) nonew (green). R=2.7, 2.1, 3.0 1x decrease. Detection with Healthy1 urine miR-21 < 1.54 HL H6914

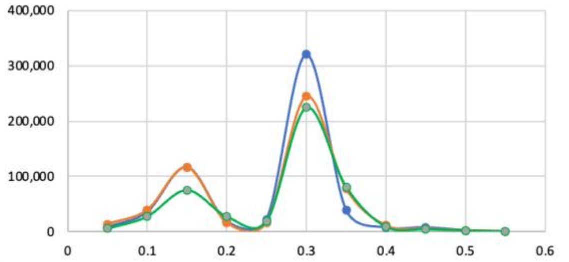

FBB12184 1.5 h: Buffer (blue) 4.4uL Healthy1 urine + 4uL probe21 (red), R=2.2, 1.9, comparable. Silencing with Healthy1 urine miR-21 > 0.70 HL H6914

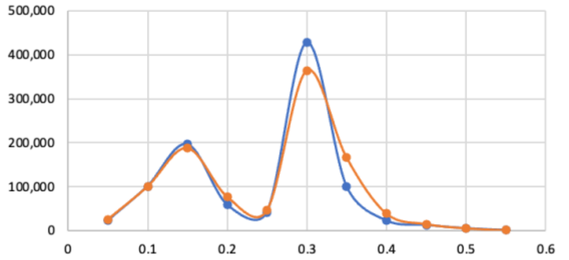

FBB10258 1.5 h: Buffer (blue) 3.7uL Healthy1 urine + 4uL probe141 (red), 3rd run 45min (green dashed) R=2.6, 2.3, 1.8, decr. Detection with Healthy1 urine miR-141 < 1.53 HL H6914

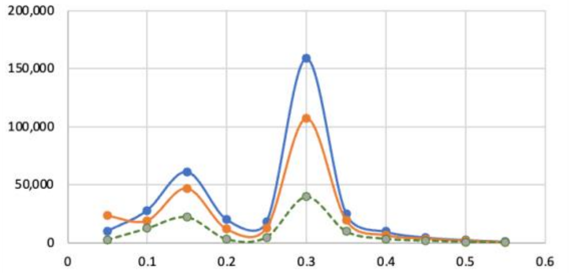

Jan21,22 FBB10383 1.5h: buffer (blue), 8uL Healthy1 urine + 4uL probe141 (red) 3rd run 45min (green dashed) R=1.0, 0.7, 0.9. Silencing with Healthy1 urine miR-141 > 0.70 HL H6914

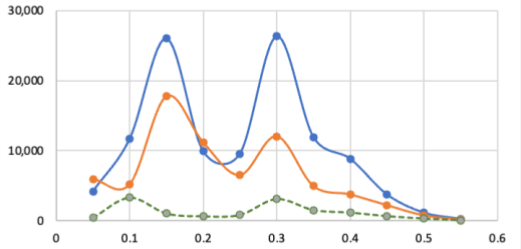

FBB10258 1.5h: Buffer (blue) 2.6uL Healthy1 urine + 4uL probe375 (red), 3rd run 45min (green, dashed), R=3.8, 3.2, 2.9. Detection with Healthy1 urine miR-375 < 1.53 HL H6914

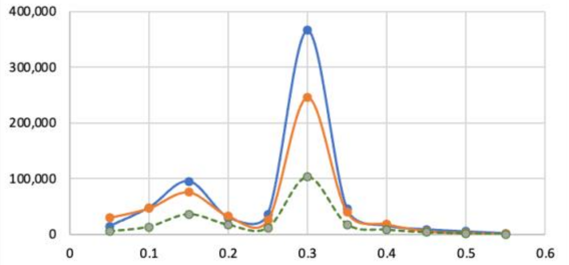

FBB10401 1.5 h: Buffer (blue), 3.6uL Healthy2 urine + 4uL probe15b (red) R=1.6, 0.9 decrease. Detection with Healthy2 urine miR15b < 1.50 HL H6914

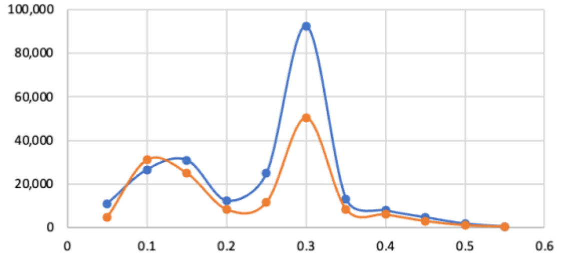

FBB11871 45min: Buffer nonew (blue) 7uL Healthy2 urine + 4uL probe15b (red), R=0.3, 0.3 comparable. Silencing with Healthy2 urine miR-15b > 0.71 HL H6914

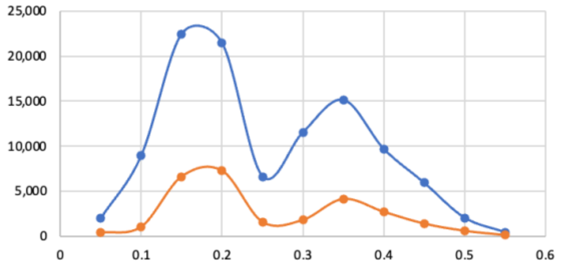

FBB11923 1.5 h: Buffer (blue) 12uL Healthy2 urine + 5uL probe21 (red) R=1.2, 1.2 comparable. Silencing with Healthy2 urine miR-21 > 0.73 HL H6914

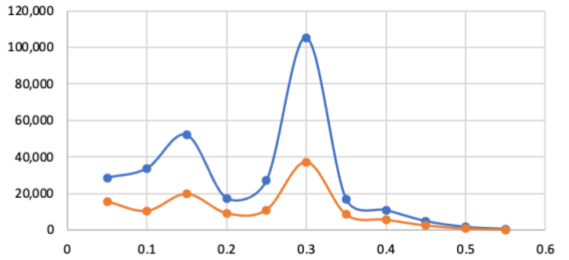

FBB00550 1.5 h: Buffer (blue) 6.4uL Healthy2 urine + 5uL probe21 (red), 45min 3rd run (green dashed), R=1.4, 1.5, 1.1. Detection with Healthy2 urine miR-21 < 1.50 HL H6914

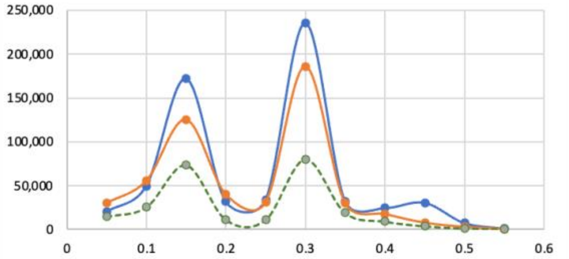

FBB28565 1.5 h: Buffer (blue) 9.3uL Healthy2 urine + 4uL probe141 (red) R=1.8, 0.5 decrease. Detection with Healthy2 urine miR-141 < 1.52 HL H6914

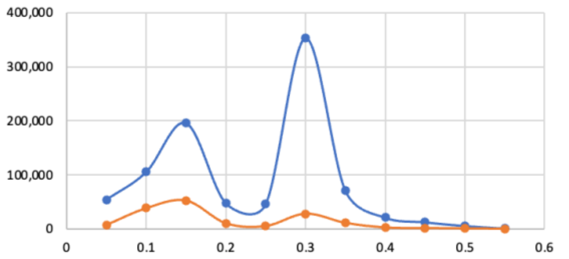

MinION/Yenos Tests rejected (not passing Quality Control); rationalization is given in the title of the figure.

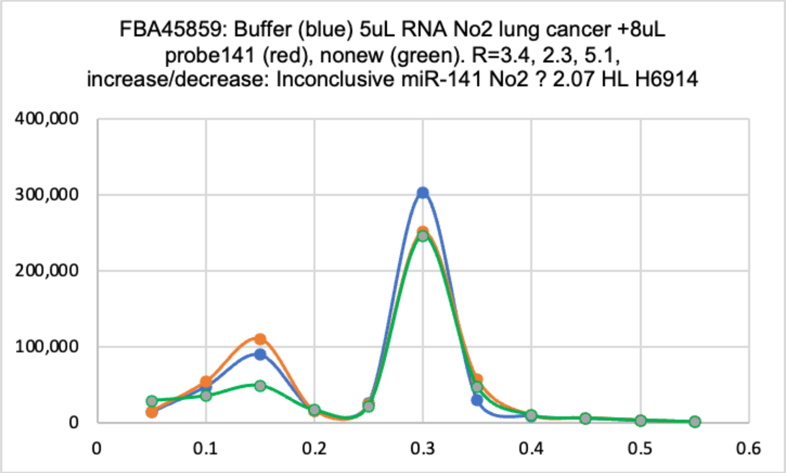


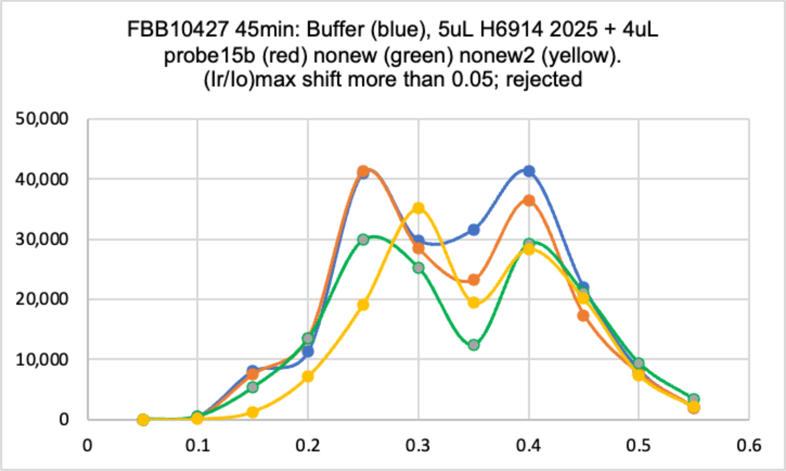


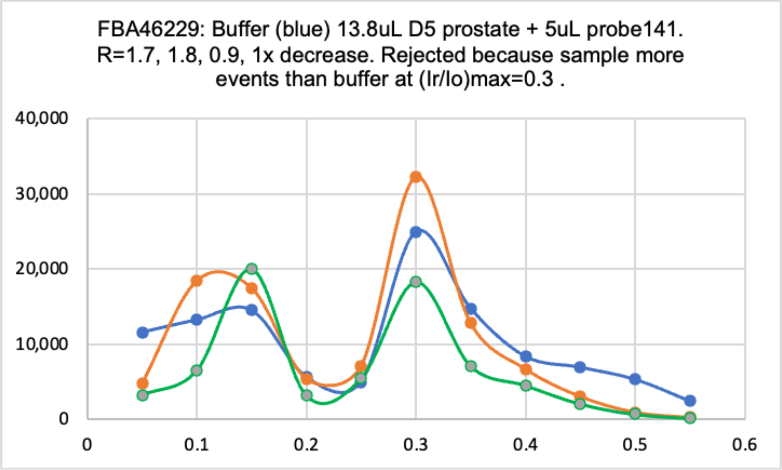
